# Brain imaging phenotypes associated with polygenic risk for Essential Tremor

**DOI:** 10.1101/2024.07.16.24310501

**Authors:** Miranda Medeiros, Alexandre Pastor-Bernier, Houman Azizi, Zoe Schmilovich, Charles-Etienne Castonguay, Peter Savadjiev, Jean-Baptiste Poline, Etienne St-Onge, Fan Zhang, Lauren J. O’Donnell, Ofer Pasternak, Yashar Zeighami, Patrick A. Dion, Alain Dagher, Guy A. Rouleau

**Affiliations:** Department of Human Genetics, McGill University, Montréal, QC, Canada; Montreal Neurological Institute-Hospital, McGill University, Montréal, QC, Canada; Department of Neurology and Neurosurgery, McGill University, Montréal, QC, Canada; Douglas Mental Health University Institute, McGill University, Montréal, QC, Canada; Université du Québec en Outaouais (UQO), Gatineau, QC, Canada; Harvard Medical School, Boston, MA, USA; School of Information and Communication Engineering, University of Electronic Science and Technology of China, Chengdu, China; Department of Psychiatry, Mass General Brigham, Harvard Medical School, Boston, MA, USA; Department of Radiology, Mass General Brigham, Harvard Medical School, Boston, MA, USA; Department of Psychiatry, McGill University, Montréal, QC, Canada

## Abstract

Essential tremor (ET) is a common movement disorder with a strong genetic basis. Magnetic resonance imaging (MRI), particularly diffusion-weighted MRI (dMRI) and T1 MRI has been used to identify brain abnormalities of ET patients. However, the mechanisms by which genetic risk affects the brain to render individuals vulnerable to ET remain unknown.

We aim to understand how ET manifests by identifying presymptomatic brain vulnerabilities driven by ET genetic risk.

We probe the vulnerability of healthy people towards ET by investigating the association of morphometry, and white and grey matter dMRI with ET in polygenic risk scores (PRS) in roughly 30,000 individuals from the UK Biobank (UKB).

Our results indicate significant effects of ET-PRS with mean diffusivity, fractional anisotropy, free water, radial diffusivity, and axial diffusivity in white matter tracts implicated in movement control. We found significant associations between ET-PRS and grey matter tissue microstructure, including the red nucleus, caudate, putamen, and motor thalamus. ET-PRS was associated with reduced grey matter volumes in several cortical and subcortical areas including the cerebellum. Identified anomalies include networks connected to surgical sites effective in ET treatment. Finally, in a secondary analysis, low PRS individuals compared to a small number of patients with ET (*N*=49) in the UKB revealed many structural differences.

Brain structural vulnerabilities in healthy people at risk of developing ET correspond to areas known to be involved in the pathology of ET. High genetic risk of ET seems to disrupt ET brain networks even in the absence of overt symptoms of ET.

## Introduction

Essential tremor (ET) is one of the most common movement disorders, affecting over 60 million people worldwide. It is characterized by action tremors predominantly in the hands, although it can also affect the legs, head, and voice. The prevalence of ET rises with age, and age at diagnosis ranges from childhood to the sixth decade.^1,2^ A family history is present in over half of patients, and twin studies demonstrate a concordance rate of 60-90% in monozygotic twins.^1^

ET is considered polygenic, with common variants explaining over 18% of the susceptibility.^3^ The underlying neural cause of ET or its genetic mechanisms remain unknown, but evidence from MRI studies and therapeutic effects of neurosurgery points to abnormalities in cerebellar-thalamic circuits.^1^

In this study, known common variants in ET were summed to generate polygenic risk scores (PRS) to quantify individual-level risk in roughly 30,000 UK Biobank (UKB) participants. PRS was combined with brain imaging phenotypes to map brain abnormalities underlying vulnerability to ET as our primary objective.^3–5^ We probed white matter (WM) dMRI, grey matter (GM) dMRI, and cortical and subcortical morphometry to show that common variant-driven ET genetic risk is associated with effects in tremor-related motor-control regions of the brain in healthy individuals. Specifically, we show that WM tissue microstructure and GM volume associations with ET-PRS implicate tracts involved in ET pathophysiology, including the thalamic ventral intermediate nucleus (VIM), cerebellum and interconnected regions that are targets of therapeutic deep brain stimulation (DBS), among others. As a secondary objective, we compare ET cases to controls of the highest and lowest genetic risk informed by ET-PRS. This identified morphometric differences between high and low-risk healthy individuals compared to ET patients in regions implicated in ET. Though the healthy individuals studied have not developed ET, our findings demonstrate that ET genetic risk affects the brain even in asymptomatic individuals. We suggest that ET develops when vulnerable tremorgenic circuitry is further damaged by as yet unknown environmental events.

## Methods

### UK biobank data

Publicly available data from the UKB consisting of genetic and MRI data from 42,488 participants was used.^4^ For data quality, we excluded individuals with poor image quality, specifically relying on the UKB processing pipeline,^6^ which identified scans with excessive head motion and artifacts, and participants with visible brain pathology. Individuals with clinical diagnoses associated with brain pathology, including ET diagnosis, and participants with non-European ancestries were excluded (eMethods). After exclusion criteria 23,552 individuals (53% female, mean age = 65.9 years, age range 49-82 years) were included in the WM analysis, 28,932 individuals (53% female, mean age = 66.0 years, age range 49-83 years) in the GM diffusion-weighted MRI analysis, and 29,706 individuals (53% female, mean age = 66.1 years, age range 49-82 years) in the cortical and subcortical volume analysis. Brain imaging was conducted on Siemens Skyra 3T running VD13A SP4 with a standard Siemens 32-channel RF receive head coil in three different centers, Manchester (2015), Newcastle, and Reading (2017).

The set of imaging-derived phenotypes (IDPs) used consisted of brain volume derivatives (from T1-weighted MRI), and dMRI derivatives (eMethods). Commonly recommended confound-regressor covariates^7^ (*acquisition date*, *age*, *age*^2^, *sex*, *age*sex*, *head motion from rfMRI* and *head motion from task fMRI*) were incorporated in all GLM (General Linear Model) statistical analyses.

### Diffusion imaging derivatives

Diffusion-weighted MRI was used to derive microstructural measures from WM and GM. WM tractograms and the standard diffusion tensor imaging (DTI) derivatives of fractional anisotropy (FA), free water (FW), mean diffusivity (MD), radial diffusivity (RD), and axial diffusivity (AD) for each UKB participant were computed using the Tractoflow^8^ pipeline (Fig. 1A-B). The pipeline performs diffusion-weighted imaging (DWI) preprocessing steps, including denoising, eddy-top-up, brain extraction, N4 Bias correction, DWI normalization, and resampling to 1mm isotropic spatial resolution (eMethods). Multiple atlases were used to limit bias and increase analysis robustness and replicability. The purpose and regions covered by atlases address (1) comprehensive anatomical regions of interest (ROI) of the entire brain with fine sampling of subcortical, brainstem, and cerebellar regions known to be involved in ET and (2) focused ROI selection based on ET surgical atlases to assess regions thought to be implicated in tremor generation (Fig 1C; eMethods)

**Figure 1.**
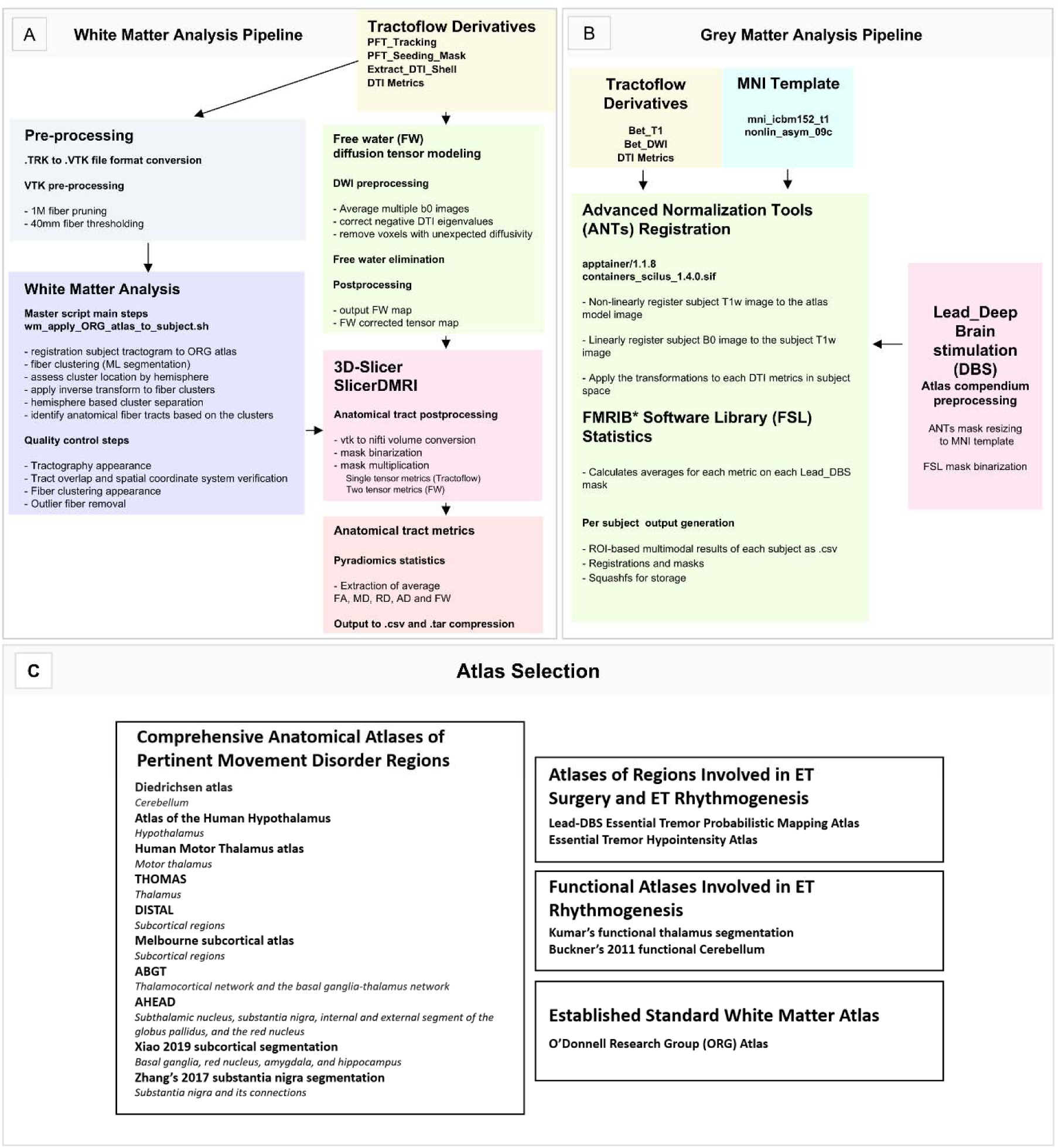
Flowcharts depicting applied methodologies. Flowchart describing the different processing steps of the white matter processing pipeline (A) and grey matter processing pipeline (B) from tractoflow derivatives. (C) Atlas selection rationale *Functional MRI Bayesian (FMRIB) (Woolrich et al. 2009)

### Polygenic risk score

Full summary statistics from the Liao *et al*.^3^ ET GWAS were used to calculate ET single nucleotide polymorphism (SNP) posterior effect estimates through PRS-CS.^5^ SNP posterior effect estimates were summed per genotype at the individual level using PLINK^9^ to generate an ET-PRS for all UKB participants after exclusion criteria.^7^

Variant quality control proceeded as previously described.^10^ Individuals with ET were removed from analysis to focus on non-symptomatic individuals.^3^ Correct ancestry groupings were verified using the 1000 Genomes Project to ensure remaining samples clustered with those of European origin to limit population stratification.^11^ Principal component analysis was done for remaining samples using PLINK to further correct for population stratification by regressing out the top 15 principal components to correct scores.^9^

### Testing associations between imaging features and ET-PRS

GLM were used to evaluate associations between imaging features (IDPs: dMRI and morphometry) and ET-PRS. The basic GLM for WM diffusion MRI analysis incorporated a mean diffusion measure (e.g. FA) as dependent variable for each tract, and the Z-score corrected ET-PRS along with the following confounding covariates (*acquisition date*, *age*, *age*^2^, *sex*, *age*sex*, *head motion from rfMRI* and *head motion from task fMRI*) as independent variables. We used ANOVA type III in R to assess statistical significance of each measure in each tract. FDR-corrected -threshold for all analyses was set at 0.05. Effect size was assessed with Eta-squared and Cohen’s F. The same approach was used for GM dMRI analysis in the Lead-DBS subcortical atlas compendium regions.

We investigated the associations between cortical surface area, cortical thickness, cortical and subcortical volume in each ROI with ET-PRS using the same GLM approach.^7^ To account for intra-cranial volume variability in cortical and subcortical segmentations, we corrected ROIs for total intracranial volume (UKB field ID: 26521-2.0).

### Comparison of genetically high-risk and low-risk individuals to ET patients

We further leveraged PRS to identify healthy controls at risk extremes. This allowed us to examine how controls at the highest and lowest risk for ET differ from UKB participants diagnosed with ET (ICD10 field 41270: diagnosis code G25.0). ET cases from the UKB were selected, ensuring all individuals were scanned under the same conditions. However, the exact diagnostic criteria used in hospitals were not provided by the UKB. In total, 49 individuals with ET diagnosis in the UKB had an MRI assessment that passed quality control. We then identified a group of control participants from the imaging cohort matched to the ET participants with R MatchIt using GLM and nearest neighbour estimation (eTable S24). Full-distance matching samples with exact ratio (1:1) were obtained with subcortical ROI volumes derived from the Diedrichsen Atlas^12^ (structural cerebellum) as the dependent variable and sex, age, scan date, and scanning center as matching covariates. Subcortical ROIs of the cerebellum were selected to expand on the extensive ET-PRS associations within these ROIs. Healthy control groups were selected according to PRS: matching subgroups were generated from the upper 5% and lower 5% quantiles (genetically high and low-risk PRS, respectively). Individuals at risk extremes were selected for this analysis as they represent either near-ideal controls (low-risk) or those at the highest risk of developing ET but are asymptomatic (high-risk) (eMethods: Fig. SS5). We used Welch t-test to assess differences between group means for each ROI as this test is more robust to sample variance differences than t-tests.^13^

### Ethics statement

UK Biobank has approval from the North-West Multi-centre Research Ethics Committee (MREC) as a Research Tissue Bank (RTB) approval and obtained patient consent for database enrolment. This research has been conducted using the UK Biobank Resource under Application Number 45551. McGill University Health Centre Research Ethics Board approved this work (Reference number: IRB00010120).

## Data Sharing

Genotyping and neuroimaging data are available through the UKB: https://www.ukbiobank.ac.uk/

Full ET summary statistics can be requested through 23andMe: https://research.23andme.com/collaborate/#dataset-access/

The code used for analysis is accessible in the following repositories, which are publicly available:

PRS-CS: https://github.com/getian107/PRScs

PLINK: http://pngu.mgh.harvard.edu/purcell/plink/

LDSC: https://github.com/bulik/ldsc

The WM dMRI code for the processing pipeline and containerized image are available in: Pastor-Bernier, Alex (2022). White matter pipeline Singularity Image

Zenodo: https://zenodo.org/records/5910831

Github: https://github.com/hayabusapb/wma_qc

The GM dMRI code for the processing pipeline is available in:

Github: https://github.com/HoumanAzizi/UKB_DTI_Pipeline

## Results

### ET-PRS associations with white matter microstructure in cerebellar and cortical tracts

ET-PRS was positively associated with MD in many WM tracts in the ORG atlas (eTable S1, Fig. S1-S2).^14^ Fig. 2 shows the associations of ET-PRS and MD in the anterior corpus callosum (CC1; *p_FDR_*=0.022), middle cerebellar peduncle (MCP; *p_FDR_*=0.021), intracerebellar input and Purkinje tract (left Intra-CBLM-I&P; *p_FDR_*=0.037), superficial frontal tract (Sup-F; *p_FDR_*=0.03), and left cortico-spinal tract (CST, *p_FDR_*=0.042). Significant associations were observed between ET-PRS and MD in the right uncinate fasciculus (UF; p_FDR_=0.038) and the left superior occipital tracts (Sup-O; p_FDR_=0.039). ET-PRS associations with FW showed similar associations to MD (eTable S1-S2), with significant FW cerebellar associations in the bilateral Intra-CBLM-I&P (*p_FDR_*≤0.002), MCP (*p_FDR_*<0.001) and the bilateral intracerebellar parallel tracts Intra-CBLM-PaT (*p_FDR_*≤0.022). Frontal, occipital, and cortico-spinal projection tracts showed significant relationships between FW and ET-PRS: left Sup-F (*p_FDR_*=0.005), bilateral Sup-O (*p_FDR_*≤0.026), and left CST (*p_FDR_*<0.042; Fig. S1-S2, eTable S2). Extended dMRI analysis (Fig. S1, eTable S3-S5) revealed that ET-PRS was positively associated with RD in the MCP (*p_FDR_*=0.014, eTable S4) and the left Intra-CBLM-I&P (*p_FDR_*=0.038), the cingulum bundle (CB; *p_FDR_*=0.015), the CC1 (*p_FDR_*=0.016), bilateral Sup-F (*p_FDR_*≤0.027), as well as the bilateral uncinate fasciculus (UF; *p_FDR_*≤0.038). Associations between ET-PRS and RD were observed in the bilateral Sup-O (*p_FDR_*<0.047) and the bilateral inferior occipitofrontal fasciculus (*p_FDR_*<0.05). ET-PRS was positively associated with AD in the striato-parietal (SP; *p_FDR_*<0.016; eTable S5), superficial-parietal-occipital (Sup-PO; *p_FDR_*<0.003), the CC1 (*p_FDR_*<0.032), and the right inferior longitudinal fasciculus (ILF; *p_FDR_*<0.037; eTable S5). We also observed negative associations of ET-PRS with FA in the left superficial frontal tract Sup-F (*p_FDR_*<0.02; eTable S3). Overall, ET-PRS was associated with increased diffusivity and free water, indicating altered tissue microstructure in several WM tracts implicated in movement control.

**Figure 2.**
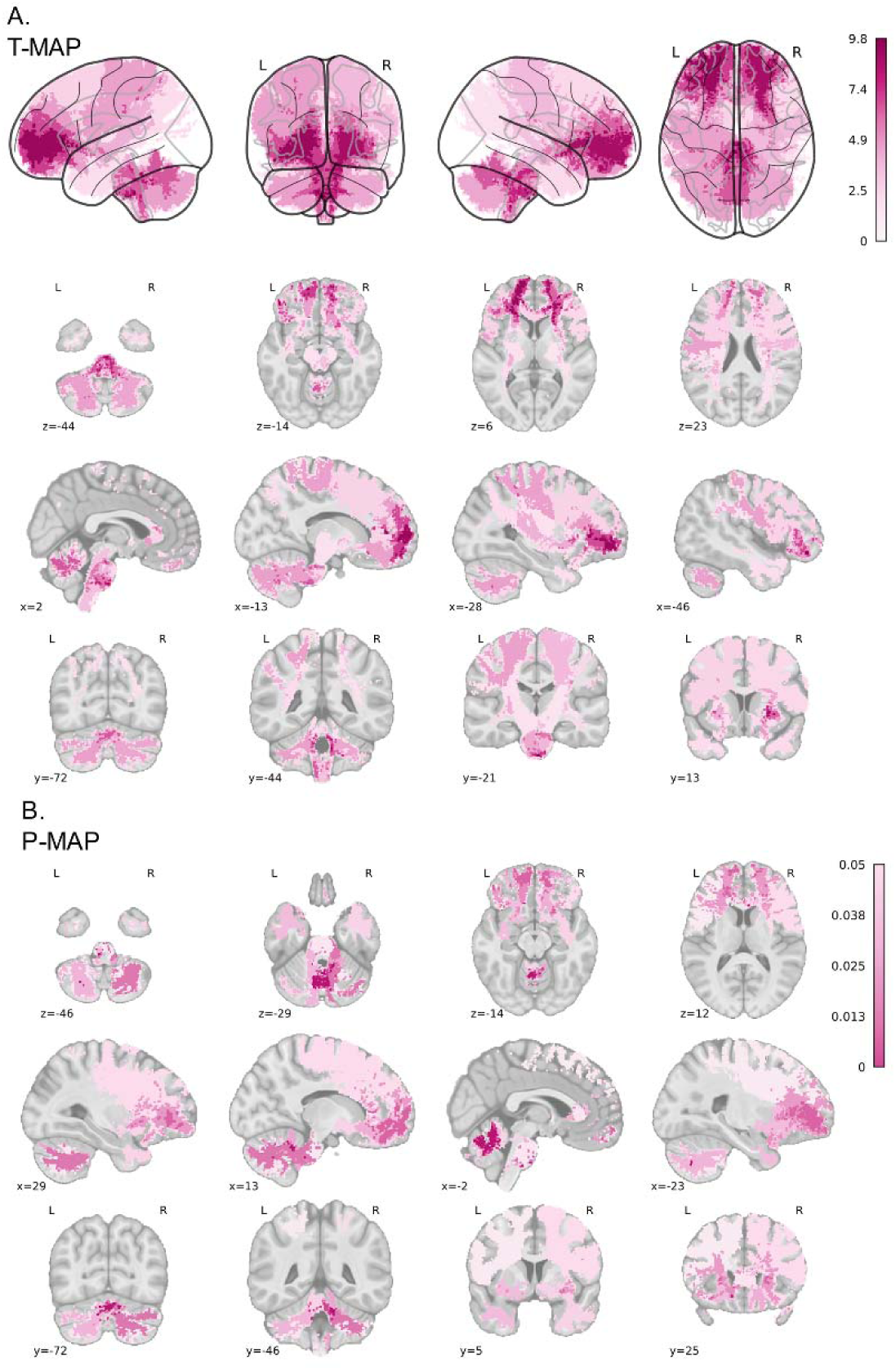
White matter diffusion-weighted magnetic resonance imaging mean diffusivity. Mean diffusivity across white matter anatomical tracts reveals positive associations with ET-PRS in cerebellar spinal and frontal projection tracts in (A) t-statistic and (B) FDR-adjusted p-value projections.^14^

### ET-PRS associations with red nucleus and thalamus grey matter microstructures

To confirm neuroanatomical findings for ET, we ran associations between dMRI measures in the GM in targets based on the Lead-DBS neurosurgical atlases (coefficients in Fig. S3, MNI_152 projections in Fig. S4-S6, all associations in eTables S6-S17). We identified regions implicated in the therapeutic effects of DBS in ET with structural and functional segmentations. We found significant positive associations of ET-PRS with FA in the red nucleus bilaterally (RN; *p_FDR_*≤0.003; eTable S6; Lead-DBS Essential Tremor Probabilistic Mapping Atlas,^15^ Essential Tremor Hypointensity Atlas,^16^ RN in the Atlas of the Human Hypothalamus,^17^ and in Xiao 2019 subcortical segmentation).^18^ We also found negative associations in Kumar’s functional thalamus segmentation in area 7 (*p_FDR_*_=_0.048; Kumar Atlas^19^; Fig. S5; eTable S10). Conversely, ET-PRS had positive associations with MD in Kumar’s functional thalamus segmentation (posterior and ventral thalamic nuclei; functional areas 3R, 4B_L, 7_L, 13A, bilateral 14, 15R; *p_FDR_*<0.034).^19^ The ET-PRS was also associated with MD in the right periventricular hypothalamic nucleus (*p_FDR_*<0.049; eTable S11) and the left hippocampus (*p_FDR_*<0.035; eTable S7, Fig. S5). The t-statistic projections in Fig. S5 show bilateral posterior and ventral thalamic nuclei associations. ET-PRS associations with RD and AD in the functional thalamic network and the hypothalamus were also observed (eTables S12-S13). No significant associations of ET-PRS with any diffusion measures for Zhang’s 2017 substantia nigra segmentation^15^ or Buckner’s 2011 functional Cerebellum^20^ were seen (eTables S6-S7 & S11).

To further refine neuroanatomical localization of these effects in the striatum, thalamus, and basal-ganglia, we investigated ET-PRS associations in the following atlases: DISTAL,^21^ Human Motor Thalamus atlas,^22^ Melbourne subcortical atlas,^23^ THOMAS,^24^ ABGT,^25^ and AHEAD.^26^ We projected the MD ET-PRS association t-statistic and adjusted MD ET-PRS association p-values on the MNI2009c non-linear template. We found significant positive associations of ET-PRS with MD throughout the thalamus (eTable S14), in particular the medial and dorsolateral nuclei (DL; Human Motor Thalamus atlas),^22^ ventral posterior (VP; Melbourne Atlas),^23^ encompassing the ventral intermediate nucleus (VIM), the dorso-intermedius externus and dorso-oralis internus nuclei (DISTAL).^21^ Left hemisphere unilateral associations with the medial dorsal, ventro-caudal, zentro-caudalis and the lateropolaris nuclei were also observed. Bilateral associations in the habenula (*p_FDR_*≤0.026; Fig. 3A) were also observed. Additionally, negative ET-PRS associations with FA were identified in the body and anterior extent of the caudate nucleus, in particular the medial caudate (mCAU), the dorsal anterior caudate (Caudate-DA) in addition to the nucleus accumbens in the Melbourne subcortical atlas (Fig. 3B).^23^ In summary, regions typically linked to ET vulnerability showed positive associations with MD and negative associations with FA, except for the red nucleus which showed a positive ET-PRS association with FA across several atlases in Lead_DBS.

**Figure 3.**
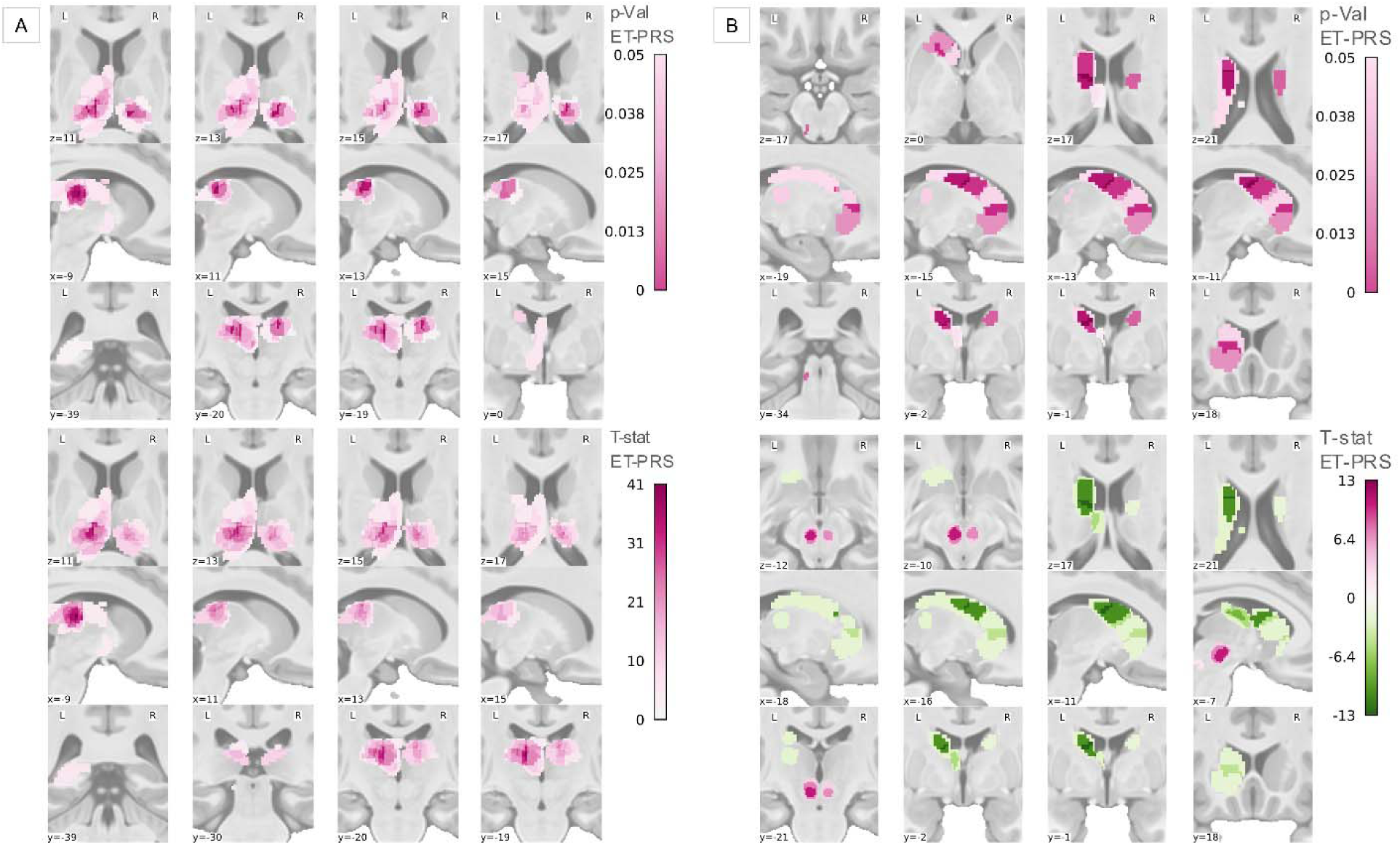
Grey matter dMRI imaging mean diffusivity and fractional anisotropy. (A) ET-PRS associations with mean diffusivity in thalamus are strongest in ventral posterior nuclei.^21,22^ (B) ET-PRS negative associations with fractional anisotropy observed in the caudate and positive associations in red nucleus.^22,23^

### ET-PRS associations with cortical surface area, cortical thickness, and cortical volume

We found limited associations of ET-PRS and cortical surface area, with positive associations in the bilateral lateral occipital (*p_FDR_*<0.037), left inferior parietal (*p_FDR_*<0.038), and right precentral area (*p_FDR_*<0.047; eTable S18). We observed many associations of ET-PRS with cortical volume (eTable S19; Fig. 4A), with negative associations in the following ROIs: bilateral superior parietal (*p_FDR_*<0.0014), bilateral caudal middle frontal (*p_FDR_*<0.021), bilateral precuneus (*p_FDR_*<0.022), left cuneus (*p_FDR_*<0.017), left posterior cingulate (*p_FDR_*<0.017), and rostral middle frontal (*p_FDR_*<0.03. In contrast, we found no associations of ET-PRS with cortical thickness, except in the right superior frontal area (*p_FDR_*<0.045; eTable S20).

**Figure 4.**
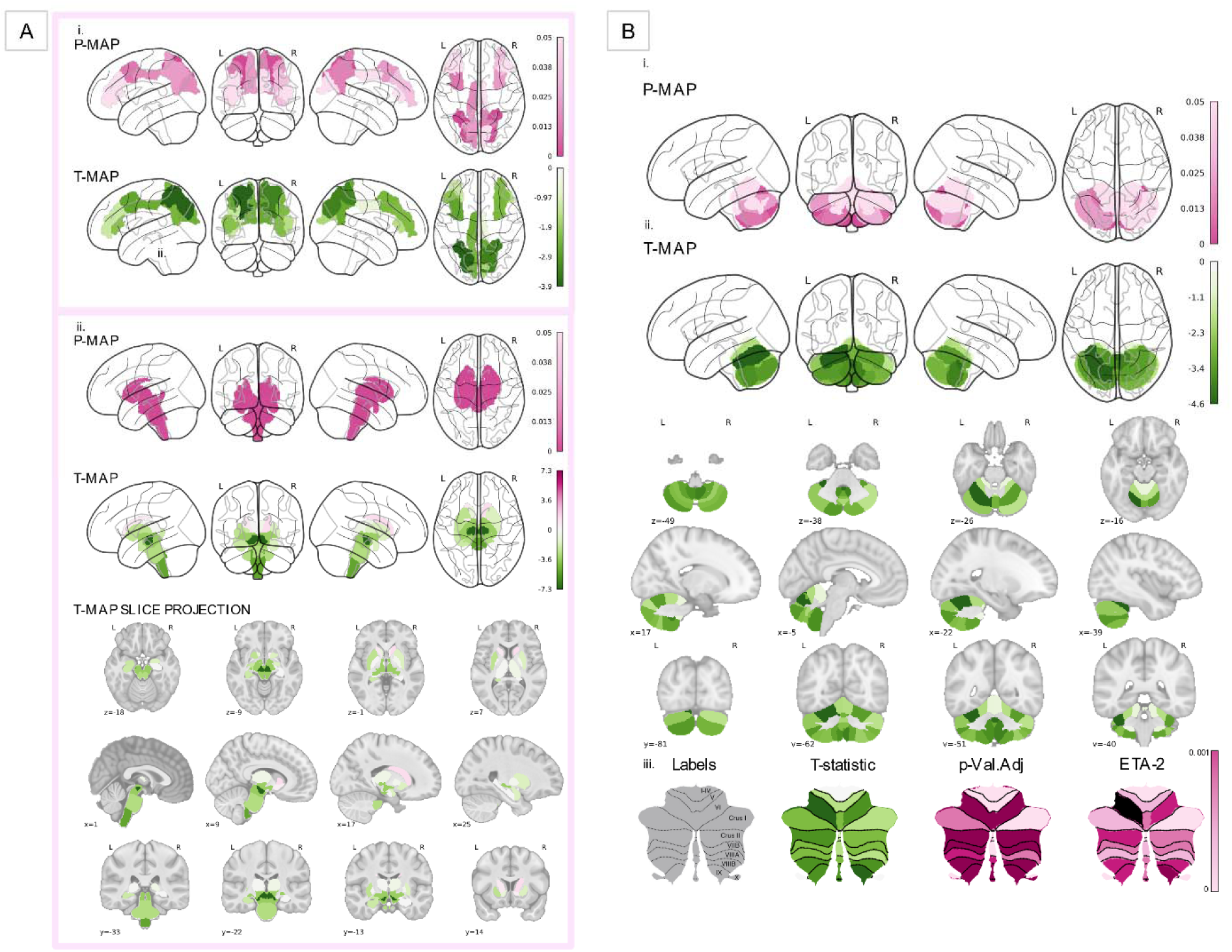
Effect of ET-PRS on grey matter volumes. (A) ET-PRS associations in the red nucleus, caudate and brainstem (i.) fractional anisotropy (ii.) p-Value projections.^23,39^ (B) Cerebellar volumes are negatively associated with ET-PRS (i.) Glassbrain p-value (ii-iii) t-statistic projections.^12^

### ET-PRS associations with brainstem and cerebellum volumes

Our analysis of UKB subcortical volume phenotypes (208 ROIs) showed negative associations with ET-PRS in the brainstem (*p_FDR_*<5.3e-4) and all its subdivisions, the midbrain (*p_FDR_*<0.001), pons (*p_FDR_*=0.0079), medulla (*p_FDR_*<0.001; eTables S21-S23), including the bilateral ventral diencephalon (ventral_DC; *p_FDR_*<0.001; eTable S23, Fig. 4A). The ventral_DC refers to a “miscellaneous” area that includes the hypothalamus, mammillary body, subthalamic nuclei, substantia nigra, red nucleus, lateral geniculate nucleus (LGN), medial geniculate nucleus (MGN), zona incerta, lenticular fasciculus, and the medial lemniscus. We also found negative ET-PRS associations with subcortical volume in the left putamen (*p_FDR_*<0.011) and the left amygdala (*p_FDR_*<0.024; eTable S22; Fig. 4A). Importantly, we found extensive negative associations of ET-PRS with cerebellar volume (eTables S22-S23). Fig. 4B shows the t-statistic projections of these associations in the left (*p_FDR_*<0.002) and right cerebellum (*p_FDR_*<0.02), including the bilateral cerebellar WM (*p_FDR_*<0.0023). We found specific associations in the following cerebellar subdivisions: left Crus I (*p_FDR_*<0.022), bilateral Crus II (*p_FDR_*<0.005), bilateral VI (*p_FDR_*<0.001), vermis VI (*p_FDR_*<0.005), bilateral VIIb (*p_FDR_*<0.022), vermis VIIb (*p_FDR_*<0.022), bilateral VIIIa (*p_FDR_*<0.029), bilateral VIIIb (*p_FDR_*<0.001), vermis of VIIIb (*p_FDR_*<0.013), bilateral IX (*p_FDR_*<0.001), vermis IX (*p_FDR_*<0.0026) and vermis of X (*p_FDR_*<0.001). We found associations in other subcortical regions like the left hippocampus (*p_FDR_*<0.03), specifically in the volume of the whole left hippocampal head (*p_FDR_*<0.045) and the molecular layer of the left hippocampal head (*p_FDR_*<0.013). Among other areas of interest in the extended subcortical segmentation, there were negative associations in the bilateral temporo-occipital division of the inferior temporal gyrus (*p_FDR_*<0.018) and the left insular cortex (*p_FDR_*<0.04; eTable S22).

### Distinct grey matter differences between ET patients and healthy controls of different genetic risk

We then compared brain morphology between diagnosed ET patients and matched control participants in the UKB (*N*=49, 1:1, eTable S24). We found lower GM subcortical volume in ET patients compared to matched genetically low-risk PRS healthy controls (Fig. 5A, Fig. S7) in the motor cerebellar regions described in eTable S25 (Diedrichsen Atlas).^12^ Lower GM volume was also found in the whole brainstem, left pallidum, right thalamus, and additional cortical regions in ET vs low-risk PRS. Higher subcortical volume between ET and low-risk PRS was found in four cortical areas: inferior division of the left lateral occipital cortex, subcallosal cortex, right orbitofrontal cortex, right pars orbitalis, and occipital fusiform gyrus (eTable S25).

**Figure 5.**
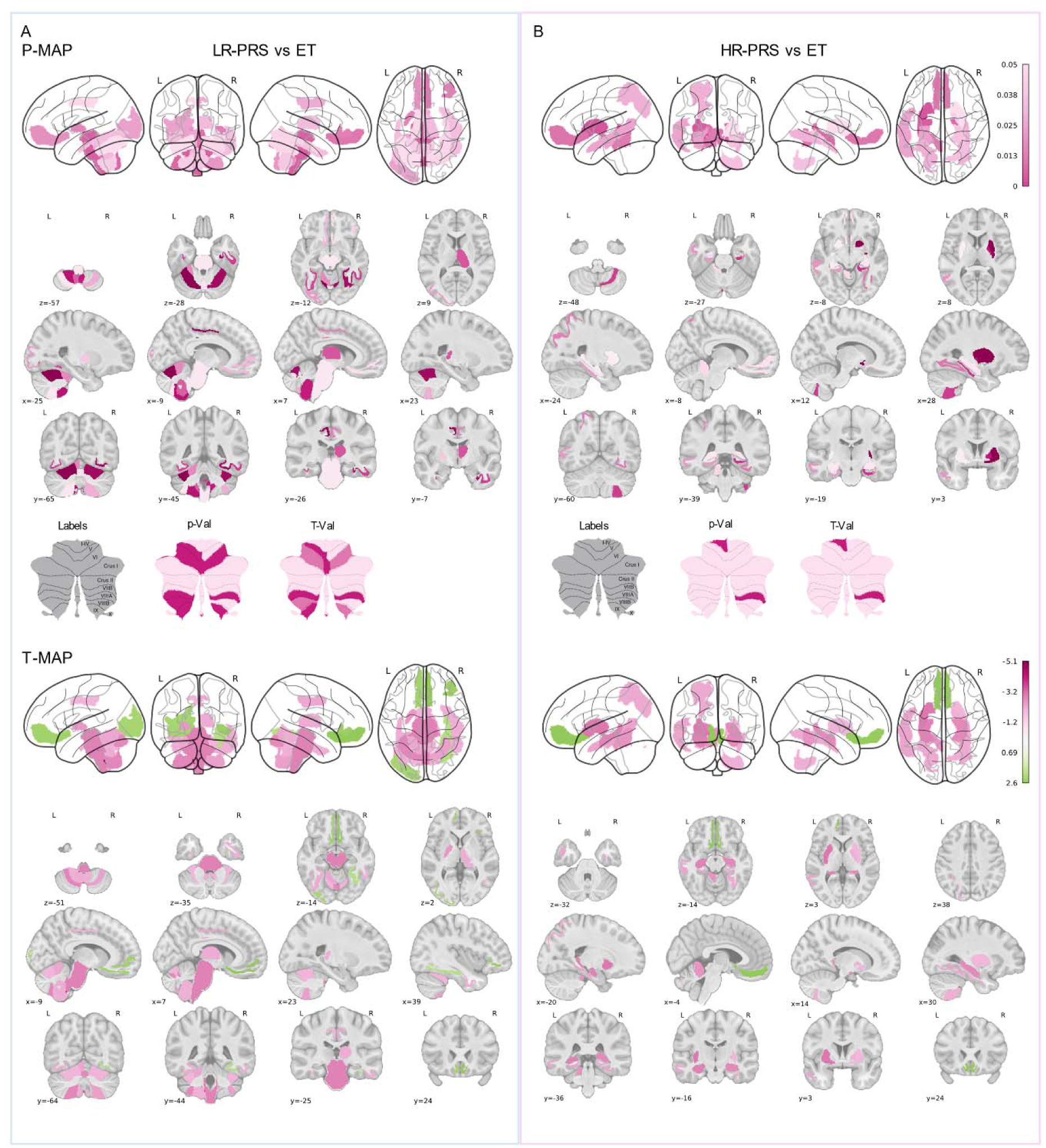
Cortical and subcortical volume differences between ET patients and controls. Differences between ET and healthy controls reveal regions associated with ET pathophysiology. Projections of t-statistics and pFDR (A) ET vs low-risk control group (B) ET vs high-risk control group.^12^

The GM volume differences between ET patients and genetically high-risk PRS healthy controls (Fig. 5B) were less extensive and unilateral in the following cerebellum areas: right VIIIa, left I2IV, and vermis Crus I (eTable S25). Lower subcortical volumes were observed in the bilateral putamen, right pallidum, and bilateral hippocampus in ET patients compared to high-risk controls (eTable S25). Lower cortical volume was also observed in ET patients compared to high-risk controls across the parahippocampal cortex, left middle temporal cortex, left superior parietal cortex, and right fusiform cortex (eTable S25). Higher cortical volume was found in the bilateral medial orbitofrontal cortex.

## Discussion

Advances in neuroimaging have revealed structural changes in regions linked to ET but have not clarified how genetic risk contributes to ET vulnerability. Our study isolates the genetic component of ET through PRS to identify WM, GM, and morphometric vulnerabilities in healthy individuals. Most regions associated with ET-PRS in this study belong to motor networks involved in tremor generation. These findings suggest that neurodevelopmental effects of ET genes may lead to subclinical changes in these circuits, thereby increasing vulnerability to environmental triggers that may bring about the full syndrome.

In our primary analysis, we looked for brain correlates of ET-PRS and identified widespread increases in MD in pathways implicated in the pathophysiology of ET, specifically in WM of the cerebellar-brainstem-thalamus-motor cortex axis. Increased MD is indicative of altered WM microstructure, reflecting increased interstitial fluid or disrupted myelination. Similar diffusion-weighted MRI alterations have been described in ET, with cerebellar and thalamic connections emerging as a consistent site of abnormality.^27–30^ The diffusion MRI findings are consistent with post-mortem structural changes in cerebellar Purkinje cells documented in ET patients.^31–33^

We also observed MD alterations in non-motor tracts, notably in orbitofrontal and prefrontal WM. Increased MD in non-motor tracts was previously reported in an ET case-control study,^28^ and may be related to subtle cognitive dysfunction,^34^ which may contribute to associations of ET with depression and anxiety.^35–38^

Associations in putative ET regions were also identified in GM, including the thalamic VIM, a common target for DBS and ablative ET therapy.^15^ ET-PRS associations with FA and MD within this area were consistently found across different atlases designed for surgical planning.^18,20–25^ We also observed associations of GM dMRI with the red nucleus across several atlases.^15,16,39^ ET-PRS was also associated with reduced GM volumes in several cortical and subcortical areas (e.g. brainstem, cerebellum, diencephalon, and putamen). This is consistent with a recent study that found significant atrophy in the middle and inferior cerebellar peduncles, and bilateral cerebellar GM in ET patients compared to controls.^40^ Additional GM associations point to regions implicated in previous studies (e.g. bilateral cerebellum, bilateral parietal lobes, right frontal lobe, and right insula), along with WM changes in the midbrain, in both occipital lobes, and right frontal lobe.^41^ Importantly, we found associations of ET-PRS with GM dMRI in various regions of the functional thalamus, red nucleus, pons, brainstem, medulla, and ventral DC which incorporate regions interconnected by the prelemniscal, cortico-pontine, and dentato-rubro-thalamic tracts in the Guillain-Mollaret triangle, a circuit coordinating cerebellar and cortical motor activity for fine motor control, where lesions may cause tremor and other movement disorders.^42^ Functional connectivity studies on patients with ET have also shown altered connectivity patterns between the cerebellum, thalamus, and several motor areas.^41^ Specifically, resting state fMRI shows abnormal functional connectivity within the cerebellar–thalamic– cortical network^43,44^ and decreased functional connectivity locally within the cerebellum in ET patients.^30,45^ The cerebellar areas associated with ET-PRS from the probabilistic tractography atlas are consistent with two distinct motor functional gradients, one located in area VI (1^st^ motor functional gradient) and a 2^nd^ motor representation covered by areas VIIB, VIIIA, VIIIB.^12^ We found the strongest associations in all three areas of the 2^nd^ motor functional gradient, particularly VIIb and VIIIa, which correspond to hand areas.^46^ These areas mainly project to the dentate nucleus, a key player in the pacemaker activity of the cerebellum involved in the cerebello-thalamo-cortical, Guillain-Mollaret triangle, and pre-lemniscal radiation pathways.^47^

Our secondary analysis focused on identifying structural differences between ET patients in the UKB cohort and controls of differing levels of genetic risk. Using the liability threshold model as our framework, PRS is used as a proxy for genetic liability. Healthy individuals with low PRS, considered near-ideal controls, carry little to no genetic vulnerability and are unlikely to develop ET. In contrast, individuals with high PRS carry greater ET genetic risk but manifest no symptoms. As expected, multiple GM volume reductions were observed in cerebellar regions of ET patients compared to low-risk individuals. Indeed, cerebellar lobule differences between cases and low-risk controls mirror many associations from the primary GM analysis: VI, VIIIa, VIIIb, IX, and X.^12^ The left V lobule is additionally affected in patients compared to low-risk controls. These findings match reports of V, VI, VIIIa, and VIIIb tremor-related activation, and decreased GM density of these lobules in patients compared to controls.^48,49^

Conversely, when comparing ET patients to high-risk controls, there were fewer differences. This suggests shared microstructural alterations between patients and high-risk individuals that may be crucial for the development of ET but are insufficient on their own to cause the disorder. However, the few differences between ET patients and high-risk controls may represent regions that must be additionally affected for ET to develop. In the cerebellum, only three regions differ between high-risk controls and ET patients: right VIIIa, left I2IV, and vermis CrusI. These regions may thus be important in the manifestation of ET. Differences between high-risk controls and patients, in other regions such as the pallidum and putamen, may also play a role in the emergence of ET. However, the VIIIa lobule of the cerebellum stands out. Here, VIIIa is different between both low-risk and high-risk controls compared to ET patients, and in other studies, showed decreased functional connectivity with the thalamus, decreased GM density, and showed tremor-related activations in patients.^48,49^ Perhaps dysfunction of the VIIIa lobule is a prerequisite for tremor onset. It should be noted, however, that the number of ET patients in the UKB was quite small. Moreover, while the diagnosis was made by participant’s clinician according to ICD-10 classification, we have no knowledge of clinical features or formal ET diagnostic criteria in these participants. Therefore, replication studies are needed.

Taken together, our findings suggest a two-hit model, where common genetic variants predispose individuals to ET, but an additional trigger, such as co-existing rare genetic variants or environmental factors like age or toxic exposures, are needed for ET to manifest.^1^ This aligns with the bimodal age of onset: familial ET cases, with higher genetic risk, show earlier onset, while sporadic cases may require more time to accumulate environmental effects and pass the threshold for disease. In our first analysis, we show that key pathways involved in the pathophysiology of ET are made vulnerable by increased genetic risk. Associations in multiple regions including the cerebellum, ventral posterior thalamus, red nucleus, and brainstem nuclei (including the pedunculo-pontine nucleus) point to a distributed network dysfunction hypothesis in ET including regions targeted by DBS ET treatment. Additionally, associations in the prefrontal and orbitofrontal areas are consistent with clinical observations of ET as a complex disorder with cognitive and psychiatric features, which may predate motor manifestation.^35–38^ It has been proposed that ET is a neurodegenerative condition, although this remains controversial.^50^ Some have suggested that aberrant oscillatory neural activity in tremor networks is the primary cause of the disease, and that MRI changes are a consequence.^50^ Our findings of microstructural alterations in relevant brain structures in association with increasing ET-PRS, in the absence of tremor, may point to a neurodegenerative etiology. In our secondary analysis, we identify the VIIIa lobule as a potential prerequisite for ET manifestation. Our findings suggest that common variants inform an endophenotype that predispose individuals to ET as additional risk factors accumulate. Future longitudinal studies of individuals of heightened genetic risk for ET could help identify the threshold where ET develops. This could lead to more effective diagnostics and therapies for this prevalent movement disorder.

## Supporting information

eMethods

supplemental assessment of environmental risk

eTables

## Data Availability

Genotyping and imaging data is available from the UK Biobank upon application.
Summary statistics of IDPs are available from the Oxford Brain Imaging Genetics Server.
ET GWAS summary statistics are available through 23andMe upon request.

https://www.ukbiobank.ac.uk/enable-your-research/apply-for-access

https://open.win.ox.ac.uk/ukbiobank/big40/

https://research.23andme.com/dataset-access/

## Acknowledgements

We are thankful to the Rouleau and Dagher lab members for their feedback and support. This work was supported by the Canadian Institutes of Health Research. We would also like to thank the employees and research participants of 23andMe, Inc. for making this work possible. This research used the NeuroHub infrastructure and was undertaken thanks in part to funding from the Canada First Research Excellence Fund, awarded through the Healthy Brains, Healthy Lives initiative at McGill University. This research was enabled in part by support provided by Calcul Québec and the Digital Research Alliance of Canada. This research has been conducted using the UK Biobank Resource under Application Number 45551.

## Authors’ Roles

(1) Analysis: A. Design, B. Execution, C. Review and Critique; (2) Manuscript Preparation: A. Writing first draft, B. Review and Critique.

MM: 1A, 1B, 1C, 2A, 2B

AP: 1A, 1B, 1C, 2A, 2B

HA: 1B

ZS: 1A, 1C, 2B

CEC: 1B, 1C, 2B

PS: 1B, 2B

JBP: 1B, 2B

ES: 1B

FZ: 1B, 2B

LJO:1B, 2B

OP:1B, 2B

YZ:1B

PAD: 1C, 2B

AD: 1A, 1C, 2B

GAR: 1A, 1C, 2B

## References

1. Welton T, Cardoso F, Carr JA, et al. Essential tremor. Nat Rev Dis Primers. 2021;7(1):83.

2. Haubenberger D, Hallett M. Essential Tremor. N Engl J Med. 2018;379(6):596–7.

3. Liao C, Castonguay CE, Heilbron K, et al. Association of Essential Tremor With Novel Risk Loci: A Genome-Wide Association Study and Meta-analysis. JAMA Neurol. 2022;79(2):185–93.

4. Sudlow C, Gallacher J, Allen N, et al. UK biobank: an open access resource for identifying the causes of a wide range of complex diseases of middle and old age. PLoS Med. 2015;12(3):e1001779.

5. Ge T, Chen CY, Ni Y, Feng YA, Smoller JW. Polygenic prediction via Bayesian regression and continuous shrinkage priors. Nat Commun. 2019;10(1):1776.

6. Alfaro-Almagro F, Jenkinson M, Bangerter NK, et al. Image processing and Quality Control for the first 10,000 brain imaging datasets from UK Biobank. Neuroimage. 2018;166:400–24.

7. Alfaro-Almagro F, McCarthy P, Afyouni S, et al. Confound modelling in UK Biobank brain imaging. Neuroimage. 2021;224:117002.

8. Theaud G, Houde JC, Bore A, Rheault F, Morency F, Descoteaux M. TractoFlow: A robust, efficient and reproducible diffusion MRI pipeline leveraging Nextflow & Singularity. Neuroimage. 2020;218:116889.

9. Chang CC, Chow CC, Tellier LC, Vattikuti S, Purcell SM, Lee JJ. Second-generation PLINK: rising to the challenge of larger and richer datasets. Gigascience. 2015;4:7.

10. Collister JA, Liu X, Clifton L. Calculating Polygenic Risk Scores (PRS) in UK Biobank: A Practical Guide for Epidemiologists. Front Genet. 2022;13:818574.

11. Genomes Project C, Auton A, Brooks LD, et al. A global reference for human genetic variation. Nature. 2015;526(7571):68–74.

12. Diedrichsen J, Balsters JH, Flavell J, Cussans E, Ramnani N. A probabilistic MR atlas of the human cerebellum. Neuroimage. 2009;46(1):39–46.

13. Ruxton GD. The unequal variance t-test is an underused alternative to Student’s t-test and the Mann–Whitney U test. Behavioral Ecology. 2006;17(4):688–90.

14. Zhang F, Wu Y, Norton I, et al. An anatomically curated fiber clustering white matter atlas for consistent white matter tract parcellation across the lifespan. Neuroimage. 2018;179:429–47.

15. Zhang Y, Larcher KM, Misic B, Dagher A. Anatomical and functional organization of the human substantia nigra and its connections. Elife. 2017;6.

16. Neudorfer C, Kroneberg D, Al-Fatly B, et al. Personalizing Deep Brain Stimulation Using Advanced Imaging Sequences. Ann Neurol. 2022;91(5):613–28.

17. Neudorfer C, Germann J, Elias GJB, Gramer R, Boutet A, Lozano AM. A high-resolution in vivo magnetic resonance imaging atlas of the human hypothalamic region. Sci Data. 2020;7(1):305.

18. Xiao Y, Lau JC, Anderson T, et al. An accurate registration of the BigBrain dataset with the MNI PD25 and ICBM152 atlases. Sci Data. 2019;6(1):210.

19. Kumar VJ, van Oort E, Scheffler K, Beckmann CF, Grodd W. Functional anatomy of the human thalamus at rest. Neuroimage. 2017;147:678–91.

20. Buckner RL, Krienen FM, Castellanos A, Diaz JC, Yeo BT. The organization of the human cerebellum estimated by intrinsic functional connectivity. J Neurophysiol. 2011;106(5):2322–45.

21. Ewert S, Plettig P, Li N, et al. Toward defining deep brain stimulation targets in MNI space: A subcortical atlas based on multimodal MRI, histology and structural connectivity. Neuroimage. 2018;170:271–82.

22. Ilinsky I, Horn A, Paul-Gilloteaux P, Gressens P, Verney C, Kultas-Ilinsky K. Human Motor Thalamus Reconstructed in 3D from Continuous Sagittal Sections with Identified Subcortical Afferent Territories. eNeuro. 2018;5(3).

23. Tian Y, Margulies DS, Breakspear M, Zalesky A. Topographic organization of the human subcortex unveiled with functional connectivity gradients. Nat Neurosci. 2020;23(11):1421–32.

24. Su JH, Thomas FT, Kasoff WS, et al. Thalamus Optimized Multi Atlas Segmentation (THOMAS): fast, fully automated segmentation of thalamic nuclei from structural MRI. Neuroimage. 2019;194:272–82.

25. He X, Chaitanya G, Asma B, et al. Disrupted basal ganglia-thalamocortical loops in focal to bilateral tonic-clonic seizures. Brain. 2020;143(1):175–90.

26. Alkemade A, Mulder MJ, Groot JM, et al. The Amsterdam Ultra-high field adult lifespan database (AHEAD): A freely available multimodal 7 Tesla submillimeter magnetic resonance imaging database. Neuroimage. 2020;221:117200.

27. Holtbernd F, Shah NJ. Imaging the Pathophysiology of Essential Tremor-A Systematic Review. Front Neurol. 2021;12:680254.

28. Nestrasil I, Svatkova A, Rudser KD, et al. White matter measures correlate with essential tremor severity-A pilot diffusion tensor imaging study. Brain Behav. 2018;8(8):e01039.

29. Shin DH, Han BS, Kim HS, Lee PH. Diffusion tensor imaging in patients with essential tremor. AJNR Am J Neuroradiol. 2008;29(1):151–3.

30. Saini J, Bagepally BS, Bhatt MD, et al. Diffusion tensor imaging: tract based spatial statistics study in essential tremor. Parkinsonism Relat Disord. 2012;18(5):477–82.

31. Babij R, Lee M, Cortes E, Vonsattel JP, Faust PL, Louis ED. Purkinje cell axonal anatomy: quantifying morphometric changes in essential tremor versus control brains. Brain. 2013;136(Pt 10):3051–61.

32. Louis ED. Essential tremor and the cerebellum. Handb Clin Neurol. 2018;155:245–58.

33. Choe M, Cortes E, Vonsattel JP, Kuo SH, Faust PL, Louis ED. Purkinje cell loss in essential tremor: Random sampling quantification and nearest neighbor analysis. Mov Disord. 2016;31(3):393–401.

34. Benito-Leon J, Mato-Abad V, Louis ED, et al. White matter microstructural changes are related to cognitive dysfunction in essential tremor. Sci Rep. 2017;7(1):2978.

35. Louis ED. Essential tremors: a family of neurodegenerative disorders? Arch Neurol. 2009;66(10):1202–8.

36. Louis ED. Non-motor symptoms in essential tremor: A review of the current data and state of the field. Parkinsonism Relat Disord. 2016;22 Suppl 1(0 1):S115–8.

37. Louis ED. Essential tremor as a neuropsychiatric disorder. J Neurol Sci. 2010;289(1-2):144–8.

38. Jhunjhunwala K, Pal PK. The Non-motor Features of Essential Tremor: A Primary Disease Feature or Just a Secondary Phenomenon? Tremor Other Hyperkinet Mov (N Y). 2014;4:255.

39. Nowacki A, Barlatey S, Al-Fatly B, et al. Probabilistic Mapping Reveals Optimal Stimulation Site in Essential Tremor. Ann Neurol. 2022;91(5):602–12.

40. Prasad S, Pandey U, Saini J, Ingalhalikar M, Pal PK. Atrophy of cerebellar peduncles in essential tremor: a machine learning-based volumetric analysis. Eur Radiol. 2019;29(12):7037–46.

41. Tikoo S, Pietracupa S, Tommasin S, et al. Functional disconnection of the dentate nucleus in essential tremor. J Neurol. 2020;267(5):1358–67.

42. Ogut E, Armagan K, Tufekci D. The Guillain-Mollaret triangle: a key player in motor coordination and control with implications for neurological disorders. Neurosurg Rev. 2023;46(1):181.

43. Popa T, Russo M, Vidailhet M, et al. Cerebellar rTMS stimulation may induce prolonged clinical benefits in essential tremor, and subjacent changes in functional connectivity: an open label trial. Brain Stimul. 2013;6(2):175–9.

44. Nicoletti V, Cecchi P, Pesaresi I, Frosini D, Cosottini M, Ceravolo R. Cerebello-thalamo-cortical network is intrinsically altered in essential tremor: evidence from a resting state functional MRI study. Sci Rep. 2020;10(1):16661.

45. Novellino F, Nicoletti G, Cherubini A, et al. Cerebellar involvement in essential tremor with and without resting tremor: A Diffusion Tensor Imaging study. Parkinsonism Relat Disord. 2016;27:61–6.

46. Saadon-Grosman N, Angeli PA, DiNicola LM, Buckner RL. A third somatomotor representation in the human cerebellum. J Neurophysiol. 2022;128(4):1051–73.

47. Matano S. Brief communication: Proportions of the ventral half of the cerebellar dentate nucleus in humans and great apes. Am J Phys Anthropol. 2001;114(2):163–5.

48. Broersma M, van der Stouwe AMM, Buijink AWG, et al. Bilateral cerebellar activation in unilaterally challenged essential tremor. Neuroimage Clin. 2016;11:1–9.

49. Dyke JP, Cameron E, Hernandez N, Dydak U, Louis ED. Gray matter density loss in essential tremor: a lobule by lobule analysis of the cerebellum. Cerebellum Ataxias. 2017;4:10.

50. Faust PL. Is essential tremor a degenerative disorder or an electric disorder? Degenerative disorder. Int Rev Neurobiol. 2022;163:65–101.

