## Supplementary material for "Brain imaging phenotypes associated with polygenic risk for Essential Tremor": eMethods

### Supplementary Methods

#### UKB cohort

This study focused on healthy individuals from the UKB with both genotyping and imaging data available. Individuals with clinical diagnoses associated with brain pathology (International Classification of Diseases, ICD-10) and participants with non-European ancestries were excluded.

Individuals with clinical diagnoses associated with brain pathology (International Classification of Diseases (ICD-10): HIV (20:24), neoplasm (69:72), mental (00:09,25,28:31,70:73,78:79), brain disorders including Alzheimer's Disease and Parkinson's Disease (00:14, 20:26, 30:32, 35:37, 40:41, 45:46, 80:83, 91:99), development disorders (00:07, 90:93, 95:99), and brain injury (04, 06:09)) and participants with non-European ancestries, based on principal component analysis (PCA) of the genotypes (UKB field ID: 21006) were excluded. Individuals with ET diagnosis (UKB data-field 20002 - Non-cancer illness code=1525) were also removed.

In our secondary analysis of healthy controls of the highest and lowest genetic risk towards ET compared to ET patients, ET patients are taken from the UKB. ET patients are those in the UKB who have undergone neuroimaging and are diagnosed with ET (ICD10 field 41270: diagnosis G25.0; Supplemental Methods). Taken from the UKB's description of the 41270 field: "this field is a summary of the distinct diagnosis codes a participant has had recorded across all their hospital inpatient records in either the primary or secondary position. Diagnoses are coded according to the International Classification of Disease version 10 (ICD-10)." By using the UKB for our ET cases, we include only participants from the same cohort, meaning same scanners and conditions, in our study to have the cleanest comparison we can.

**White Matter:** After exclusion criteria 23,552 individuals (53% female (n=12,468), mean age = 65.9 years, age range 49-82 years) were included in the WM diffusion-weighted MRI analysis:

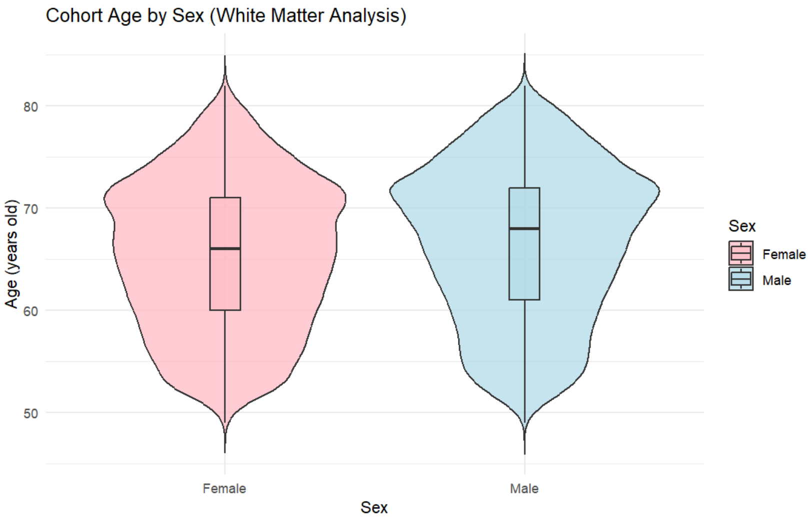

**Grey Matter:** 28,932 individuals (53% female (n=15,226), mean age = 66.0 years, age range 49-83 years) in the GM diffusion-weighted MRI analysis:

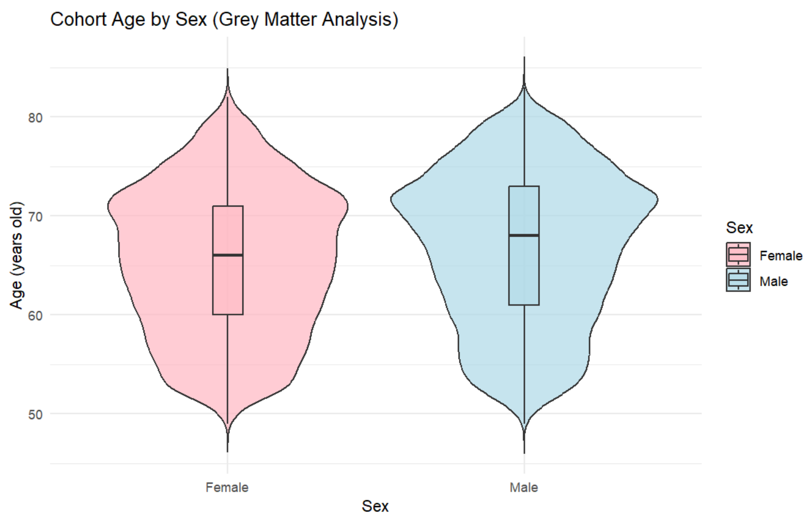


**Cortical and Subcortical:** 29,706 individuals (53% female (n=15,598), mean age = 66.1 years, age range 49-82 years) in the cortical and subcortical volume analysis:

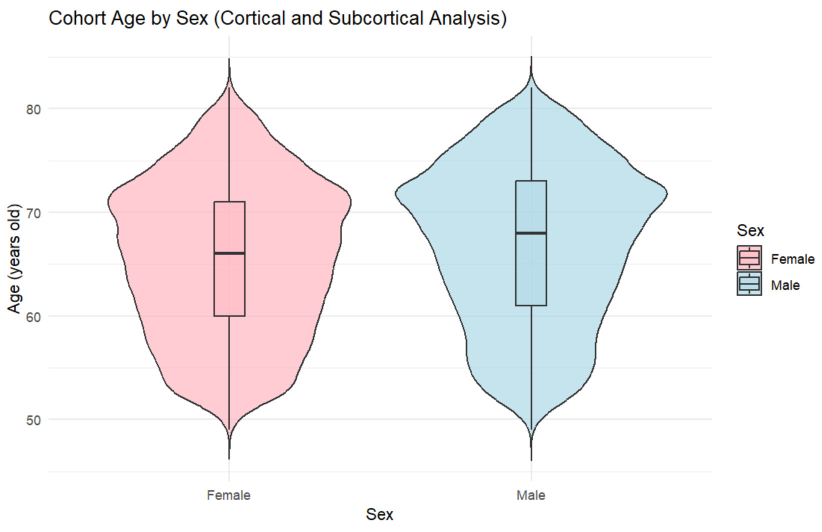


### Imaging derived phenotypes

The set of imaging derived phenotypes (IDPs) used consisted of brain volume derivatives (from T1-weighted MRI), and dMRI derivatives. IDPs were either calculated and quality-controlled by the UKB and the Centre for Functional MRI of the Brain at the University of Oxford, or by the authors of the present manuscript as described below. T1-weighted structural images (defaced) were used to obtain brain masks using BET (Brain Extraction Tool)[1] and FLIRT (Functional Magnetic Resonance Imaging of the Brain (FMRIB)'s Linear Image Registration Tool),[2,3] was used to register the masked brain to the Montreal Neurological Institute (MNI) 152 2006 non-linear standard-space template. FAST (FMRIB's Automated Segmentation Tool)[4] was used for tissue-type segmentation. FAST was used to generate 139 IDPs, by summing the GM partial volume estimates within 139 regions of interest (ROIs). These ROIs are defined in MNI152 space, combining parcellations from several atlases: the Harvard-Oxford cortical and subcortical atlases,[5-8] and the Diedrichsen cerebellar atlas.[9] FreeSurfer modelling v6.0.0 was used to estimate cortical surface.[10-12] Surface atlases were used to extract IDPs of regional cortical surface area, volume and thickness resulting in 66 ROIs.[7-13] Subcortical regions were extracted using FreeSurfer's aseg tool, and further segmentation of subcortical regions was carried out.[14]

### Scanning sequences

Brain imaging was conducted on Siemens Skyra 3T running VD13A SP4 with a standard Siemens 32-channel RF receive head coil in 3 different centers, Manchester (2015), Newcastle, and Reading (2017). For T1w imaging, 3D MPRAGE, sagittal, in-plane acceleration iPAT=2, prescan-normalise as used. The resolution was 1mm isotropic with a 208x256x256 field-of-view matrix. The duration of the scan was 5 minutes. For diffusion imaging, the resolution was 2mm isotropic, with a 104x104x72 field-of-view matrix. The duration of the scans were 7 minutes, including 36 seconds for phase-encoding reversed data. Five diffusion directions were acquired at b=0 (+3 blip-reversed), and 50 diffusion directions were acquired for two diffusion-weighted shells (b=1000 and b=2000 mm/s2, total of 100 distinct diffusion directions). The gradient timings were $\delta$=21.4 ms, $\Delta$=45.5 ms; Spoiler b-value = 3.3 s/mm 2. SE-EPI with x3 multislice acceleration, no iPAT, fat saturation. The diffusion preparation was a standard ("monopolar") Stejskal-Tanner pulse sequence.

### Diffusion weighted derivatives – Tractoflow specifics

Diffusion-weighted MRI was used to derive microstructural measures from white and grey matter. White matter tractograms and the standard diffusion tensor imaging (DTI) derivatives of fractional anisotropy (FA), mean diffusivity (MD), radial diffusivity (RD), and axial diffusivity (AD) for each UKB participant were computed using the Tractoflow[15] pipeline (Fig. 1A & 1B). The pipeline performs diffusion weighted imaging (DWI) preprocessing steps, including denoising, eddy-top-up, brain extraction, N4 Bias correction, DWI normalization, and resampling to 1mm isotropic spatial resolution. Standard diffusion derivatives are then obtained from dMRI modeling (only b-values under 1,200 mm/s). In parallel, T1 preprocessing, DWI registration (b0), and tissue segmentation are obtained in order to produce inclusion, exclusion and seeding masks for tractography. Tracking then proceeds using the fiber orientation distribution function metrics (fODF) image, the obtained inclusion and exclusion maps, and the white matter seeding mask. The tracking algorithm used was constrained particle filter tracking and a probabilistic method with ten

(10) seeds per voxel from the white-matter mask.[16] In addition, free water (FW) was computed in the white matter skeleton (pft_seeding_mask.nii.gz) from Tractoflow[15] output. A Matlab script, FreeWater_OneCase.m was used to perform preprocessing, free water elimination and postprocessing as specified by Pasternak et al.[17] FW is defined as the compartment of water molecules that do not experience flow or diffusion restriction. In the FW model two compartments are represented by two tensors corresponding to a tissue and a water compartment. The tissue compartment follows DTI's formalism.[17,18] The water compartment is modeled by an isotropic tensor with fixed diffusivity of water in body temperature as previously described.[17]

WM tractograms using 1M streamlines and 4mm fiber-length threshold for each subject were then registered to the O'Donnell Research Group (ORG) anatomically curated atlas.[19] Registration, tract segmentation and average diffusion measure extraction for each tract was obtained via a WM processing pipeline which uses WM analysis machine learning for tract segmentation (https://github.com/hayabusapb/wma_qc).[20] ORG atlas segmentation yielded 800 fiber clusters organized into 73 anatomical tracts, including 58 deep WM tracts, major long-range association and projection tracts, commissural tracts, and tracts related to the brainstem and cerebellar connections.

GM diffusion weighted derivatives were computed for subcortical structures. To identify relevant subcortical structures, we used the Lead_DBS toolbox, which contains subcortical and WM atlases used in planning deep brain surgery for neurological conditions such as ET.[21] A compendium atlas of subcortical ROIs was generated and applied to the processed DWI data with a GM processing pipeline (https://github.com/HoumanAzizi/UKB_DTI_Pipeline). ROI masks were resized, binarized and registered from atlas space (*ICBM 2009b* non-lin asym.) to subject space as described in Fig. 1. Tractoflow[15] diffusion measures masks in the MNI space were multiplied to each mask and an average FA, MD, RD and AD measure obtained for each ROI.

#### Cortical and subcortical Freesurfer segmentation

FreeSurfer modelling v6.0.0 was used to estimate cortical surface.[10,11,13] Surface atlases were used to extract IDPs of regional cortical surface area, volume and thickness resulting in 66 ROIs.[7-13] Subcortical regions were extracted using FreeSurfer's aseg tool, and further segmentation of subcortical regions was carried out.[14] Freesurfer subcortical volumetric measures were obtained for subcortical regions in addition to global brain volume, grey matter, white matter and cerebrospinal fluid (CSF) volumes corresponding to the following UKB IDs: 30710, 26514, 26518, and 26527. 208 ROIs were obtained in the process. All FreeSurfer outputs were quality control checked with the Qoala-T approach by the UKB.[22]

These 208 ROIs come from combining the probabilistic atlases covering cortical and subcortical structural areas, derived from structural data and segmentations kindly provided by the Harvard center for morphometric analysis, the Harvard- Oxford cortical and subcortical atlases, the

Diedrichsen cerebellar atlas and standard Freesurfer outputs. The use of ~200 ROIs is a reasonable compromise between too few regions and voxel-based analyses (that lose power). Combining multiple atlases has been shown to reduce bias. The rationale for the atlases used (i.e. their relevance to tremor) is in the Atlas Selection section below.

These atlases are incorporated into standard FSL packages and are widely employed.

#### Subcortical Atlases

To identify relevant subcortical structures, we used the Lead_DBS toolbox, which contains subcortical and white matter atlases used in planning deep brain surgery for neurological conditions such as ET.[21] A compendium atlas of subcortical ROIs was generated and applied to the processed DWI data with a grey matter processing pipeline (https://github.com/HoumanAzizi/UKB_DTI_Pipeline). ROI masks were resized, binarized and registered from atlas space (*ICBM 2009b* non-lin asym.) to subject space as described in Fig. 1. The atlases used to generate the subcortical ROIs were: Nigral organization atlas,[23] Essential Tremor Probabilistic Mapping,[24] Essential Tremor Hypointensity,[25] DBS targets,[26] DBS Tractography Atlas,[27] Dystonia Response Tract Atlas,[28] Cerebellar Functional Networks,[29] Atlas of the Human Hypothalamus,[30] TOR-signPD,[31] Thalamic Functional Atlas,[32] TOR-PSM dystonia,[33] Striatal Functional Networks,[34] Brainstem Connectome Atlas,[35] the Zona Incerta Atlas,[36] DISTAL (DBS intrinsic template Atlas),[37] Human Motor Thalamus atlas,[38] Melbourne subcortical atlas,[39] THOMAS (Thalamus optimized multi-atlas segmentation),[40] ABGT (Atlas of Basal Ganglia and Thalamus),[41] AHEAD (Amsterdam Ultra-high field adult lifespan database),[42] HybraPD Atlas,[43] and Xiao's subcortical atlas.[44] Grey matter masks were obtained from each atlas in Lead-DBS. All masks were binarized and registered to the MNI 152 2009c non-linear template. Tractoflow[15] diffusion measures masks in the MNI space were multiplied to each mask and an average FA, MD, RD and AD measure obtained for each ROI.

#### Atlas Selection

None of the atlases presented here were selected randomly, nor did they consist of unsuitable age range cohorts or present incompatible specific sex or ancestry groups. The criteria for atlas selection can be summarized as follows. The atlases were selected for their pertinence to the motor disorder we are studying. For instance, Diedrichsen[9] and Buckner[29] structural and functional atlases were included because of the known involvement of cerebellar structures in ET rythmogenesis. Various atlases of the thalamus Lead-DBS compendium such as Lead-DBS Essential Tremor Probabilistic Mapping Atlas[24] and Essential Tremor Hypointensity Atlas[25] were included in order to probe regions that are typically suitable targets for ET stimulation or are involved in ET rhythmogenesis. Other atlases like the atlas of Atlas of the Human Hypothalamus,[30] DISTAL,[37] Ilinsky's 2018 Human Motor Thalamus Atlas,[38] Melbourne Subcortical Atlas,[39] THOMAS,[40] ABGT,[41] and AHEAD[42] are well established reference atlases that provide an overlap with the ROI that more specific atlases provide. Their inclusion goes along the rationale of that multiple atlases can provide robustness in brain-phenotype association studies.[45] A set of atlases such as MNI-PD25[44] or the SN atlas[23] provide regions implicated in broader movement disorders. Finally, a set of atlases based on functional connectivity (subcortical,[38] Tian 2020, cerebellum,[29] human thalamus Kumar[31]) targeting ROIs involved in ET rythmogenesis were incorporated to complement anatomical atlases with a functional scope in mind.

The ORG white matter atlas[19] was chosen the analysis of white matter diffusion.

### Confounding variables

Commonly recommended confound-regressor covariates[46] were incorporated in all GLM (General Linear Model) statistical analyses used in this study: acquisition date (UKB field ID: 21862), age (UKB field ID: 21003), age$^2$, sex (UKB field ID: 31), age*sex, head motion from rfMRI (UKB field ID: 25741), and head motion from task fMRI (UKB field ID: 25742).

### Sensitivity

In our study, the inclusion of common covariates such as age and sex are standard practice aimed at aligning our results with previous research, rather than exploring how regressions are influenced by the number of regressors and the inclusion of interaction terms like age*sex, we adhered to current literature standards. When addressing sensitivity concerning atlases, it's noteworthy that we used multiple atlases, each with its own focus—some on basal ganglia, others on hypothalamus, for instance—which overlap in regions like the red nucleus and substantia nigra. This diversity in atlases allowed us to identify significant associations in each region separately. We tested sensitivity of the ET-PRS associations with imaging phenotypes (cortical and subcortical regions dMRI and morphometry) across various Lead-DBS atlases, which provide alternative segmentations on regions that are normally studied in ET. This ensured a level of robustness that a single-atlas approach could not provide.

### Polygenic Risk Score Calculations

ET PRS was calculated for all individuals with both genotyping and MRI data in the 2019 release of UKB, yielding a sample size of 29,706 (white and grey matter diffusion-weighted MRI analysis) and 30,426 (cortical and subcortical volume analysis) individuals after application of previously mentioned exclusion criteria.[46] At the variant level, quality control of the UKB genotyped samples consisted of removing: ambiguous variants (palindromic SNPs with effect allele frequencies between 0.4 and 0.6), variants with imputation information less than 0.4, variants with minor allele frequency less than 0.005, variants that fail Hardy-Weinberg Equilibrium (1×10-5), variants with missingness greater than 0.02, and variants with more than 2 alleles.[47]

### Assessment of Lesion Presence:

To address the concern that difference may be attributable to possible unknown vascular disease in the comparison of ET cases and to high-risk and low-risk controls, the load of vascular brain lesions was compared between groups. We obtained total white matter hyperintensity distributions (from T1 and T2_FLAIR images) in ET, HR-PRS and LR-PRS patients from the UKB imaging group (supplied as an image-derived phenotype by UKB). There is no statistically significant difference (W-test) between them: ET vs LR (p=0.89) and ET vs HR (p=0.76), suggesting that vascular lesions do not differ between groups.

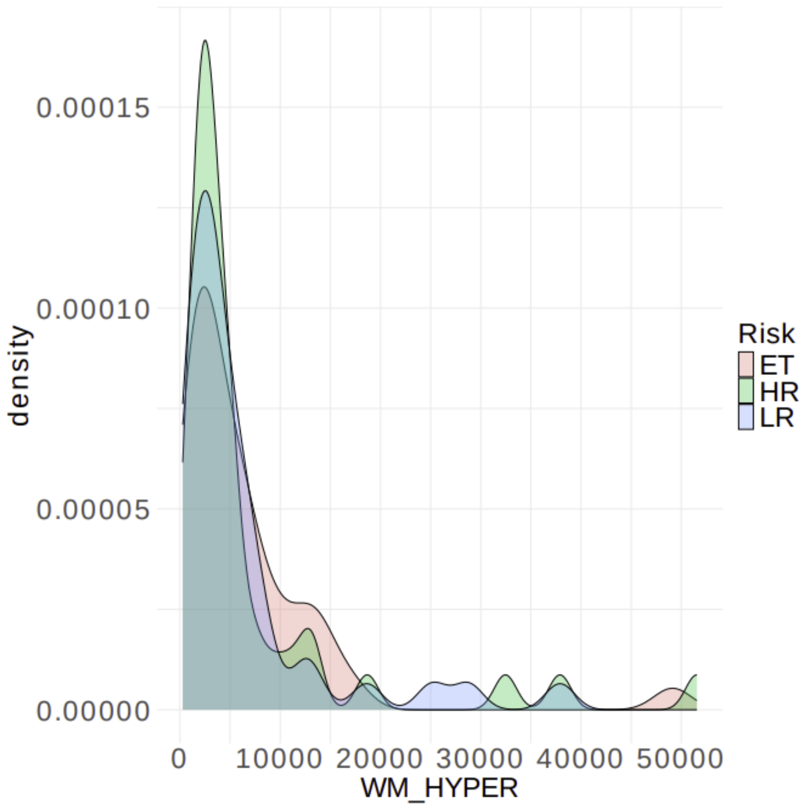


**Figure SS1. Hyperintensity profiles in matched participants (ET vs HC PRS) in the UKB cohort.** Plot of T1w whole gm. hyperintensity distributions across the set of matched subjects (#49) in ET patients (pink) vs high-risk healthy controls (green) and low-risk healthy controls (blue). No statistical differences were found for hyperintensity across the three groups with a Welch two tailed t-test: LR vs ET (t=-0.13, pVal=0.90), HR vs ET (t=0.3, p=0.76), LR vs HR (t=0.43, p=0.67)

#### Limitations

Some limitations include the generalizability of the PRS. The PRS was based on a GWAS of only European samples, so it does not hold in non-european populations. Through associating MRI measures with PRS as a proxy for genetic risk, we can only examine the consequence of SNPs which were included in the GWAS, which means other larger or rarer variant types such as structural copy number variants or repeat expansions are not analysed. As datasets expand, and as more diverse genomes are studied, these limitations will eventually be diminished.

Another limitation of this study is that it is cross-sectional in nature, limiting our ability to assess developmental or neurodegenerative trajectories related to ET PRS. Also, there are a number of ET phenotypic variants that may include different pathophysiology. Indeed, cerebellar volume differences between healthy controls and ET patients have been shown to differ substantially between different motor variants, with patients manifesting head tremors having greater differences in cerebellar volume versus healthy controls than ET patients manifesting with arm-tremors.

Finally, in the analysis comparing ET patients to healthy controls of high and low genetic risk for ET, the sample size of patients are rather small. This creates a limitation of power and generalizability.

#### Polygenic Risk Scores

The distribution of polygenic risk scores is as follows in the UKB samples:

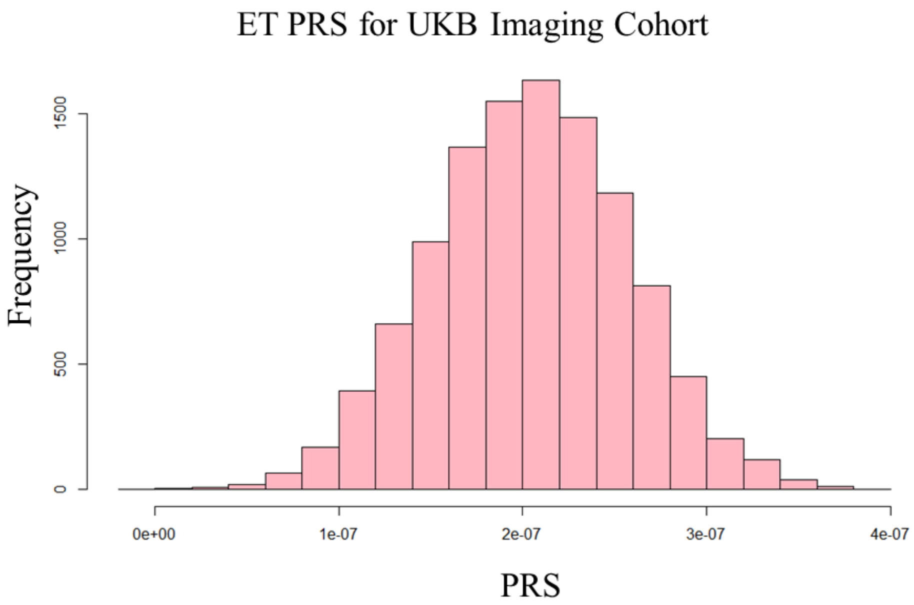


**Figure SS2. Polygenic risk score distribution.** Distribution of ET polygenic risk scores calculated in UK Biobank (UKB) cohort.

The ancestry of all samples were verified as European:

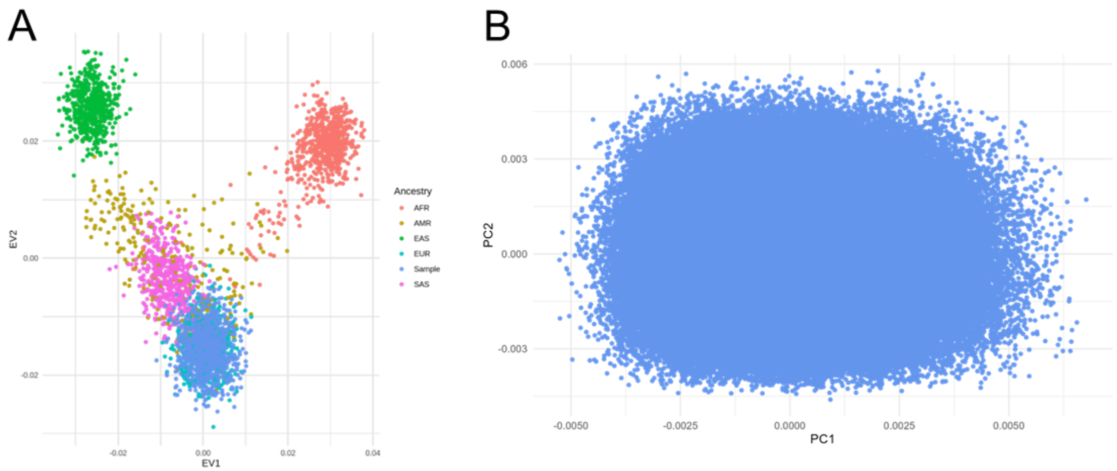


**Figure SS3. Ancestry principal component analysis.** Genetic principal component analysis. (A) The first and second principal components of a random subset of 1000 samples from the cohort is taken and is compared to 1000 genomes reference panel. Samples overlap with references of European ancestry, showing that only samples of European ancestry were considered in the analysis. (B) The first and second principal components of all samples of the cohort are depicted, the lack of any substructures suggests individuals are of a similar genetic background which reduces the risk of confounding due to subpopulations or stratification.

The screeplot which corresponds to the calculated principal components (1-15) are depicted below:

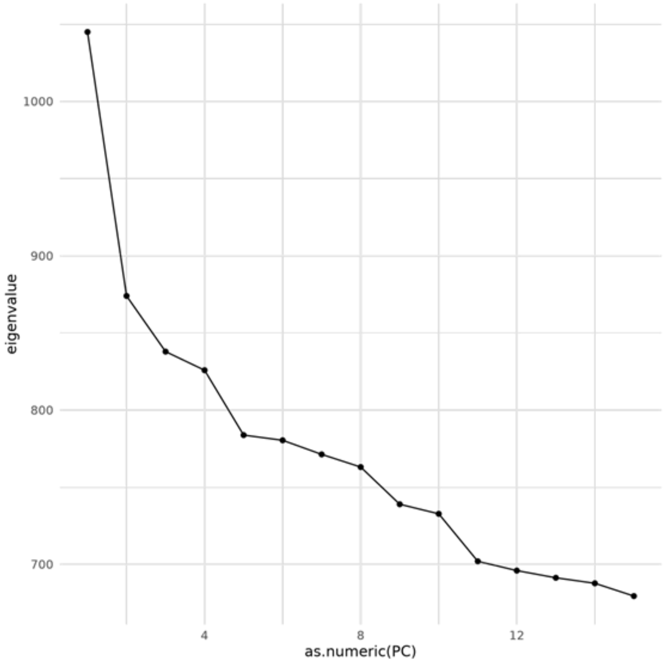


**Figure SS4. Principal component screeplot.** Scree plot of top 15 genetic principal components of cohort. Polygenic scores were then corrected by regressing out the top 15 principal components.

#### High Risk and Low Risk Controls Compared to Patients

In the analysis comparing matched patients to controls with the largest PRS (greatest genetic risk for ET) and to the smallest PRS (least genetic risk for ET), the framework is based on the threshold liability model. This model depicts risk as a normal distribution across a population where there exists some threshold which when passed, causes the onset of disease. As we are looking at healthy controls, we can assume that these individuals are below that threshold. We compare these healthy individuals of the lowest and the greatest extremes of genetic risk to ET patients to see what might be required to push individuals past the threshold to then develop disease. The following diagram depicts this framework:

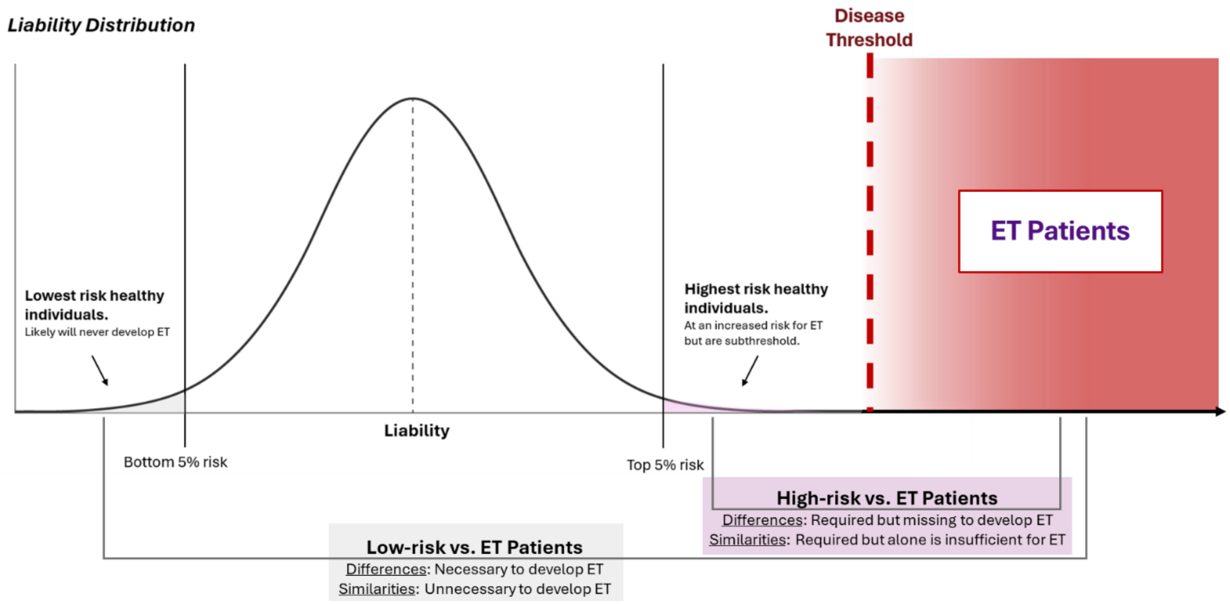


**Figure SS5. Framework of ET genetic low and high risk healthy individual groups compared to ET patients analysis through liability distribution of risk model.** The liability distribution of a healthy population's liability towards developing ET remains below disease threshold. However, as risk accumulates, healthy individuals shift towards and may even surpass the disease threshold and develop ET. Healthy individuals with the lowest polygenic risk scores (PRS) are in the lowest risk group and will likely never develop ET. On the other side of the distribution, healthy individuals with the largest PRS are in the highest risk group and are at the greatest risk of developing ET yet remain subthreshold for disease. This model provides the framework for comparisons of brain morphometry between the genetically low-risk group and patients, and comparisons between the genetically high-risk group and patients provide unique insights into what vulnerability regions are necessary for ET development.

#### References


1. Smith SM. Fast robust automated brain extraction. Hum Brain Mapp. 2002;17(3):143-55.
2. Jenkinson M, Smith S. A global optimisation method for robust affine registration of brain images. Med Image Anal. 2001;5(2):143-56.
3. Jenkinson M, Bannister P, Brady M, Smith S. Improved optimization for the robust and accurate linear registration and motion correction of brain images. Neuroimage. 2002;17(2):825-41.
4. Zhang Y, Brady M, Smith S. Segmentation of brain MR images through a hidden Markov random field model and the expectation-maximization algorithm. IEEE Trans Med Imaging. 2001;20(1):45-57.
5. Makris N, Goldstein JM, Kennedy D, et al. Decreased volume of left and total anterior insular lobule in schizophrenia. Schizophr Res. 2006;83(2-3):155-71.
6. Frazier JA, Chiu S, Breeze JL, et al. Structural brain magnetic resonance imaging of limbic and thalamic volumes in pediatric bipolar disorder. Am J Psychiatry. 2005;162(7):1256-65.
7. Desikan RS, Segonne F, Fischl B, et al. An automated labeling system for subdividing the human cerebral cortex on MRI scans into gyral based regions of interest. Neuroimage. 2006;31(3):968-80.
8. Goldstein JM, Seidman LJ, Makris N, et al. Hypothalamic abnormalities in schizophrenia: sex effects and genetic vulnerability. Biol Psychiatry. 2007;61(8):935-45.
9. Diedrichsen J, Balsters JH, Flavell J, Cussans E, Ramnani N. A probabilistic MR atlas of the human cerebellum. Neuroimage. 2009;46(1):39-46.
10. Dale AM, Fischl B, Sereno MI. Cortical surface-based analysis. I. Segmentation and surface reconstruction. Neuroimage. 1999;9(2):179-94.
11. Fischl B, Sereno MI, Tootell RB, Dale AM. High-resolution intersubject averaging and a coordinate system for the cortical surface. Hum Brain Mapp. 1999;8(4):272-84.
12. Fischl B, van der Kouwe A, Destrieux C, et al. Automatically parcellating the human cerebral cortex. Cereb Cortex. 2004;14(1):11-22.
13. Fischl B, Sereno MI, Dale AM. Cortical surface-based analysis. II: Inflation, flattening, and a surface-based coordinate system. Neuroimage. 1999;9(2):195-207.
14. Iglesias JE, Augustinack JC, Nguyen K, et al. A computational atlas of the hippocampal formation using ex vivo, ultra-high resolution MRI: Application to adaptive segmentation of in vivo MRI. Neuroimage. 2015;115:117-37.
15. Theaud G, Houde JC, Bore A, Rheault F, Morency F, Descoteaux M. TractoFlow: A robust, efficient and reproducible diffusion MRI pipeline leveraging Nextflow & Singularity. Neuroimage. 2020;218:116889.
16. Girard G, Whittingstall K, Deriche R, Descoteaux M. Towards quantitative connectivity analysis: reducing tractography biases. Neuroimage. 2014;98:266-78.
17. Pasternak O, Sochen N, Gur Y, Intrator N, Assaf Y. Free water elimination and mapping from diffusion MRI. Magn Reson Med. 2009;62(3):717-30.
18. Basser PJ, Pierpaoli C. Microstructural and physiological features of tissues elucidated by quantitative-diffusion-tensor MRI. J Magn Reson B. 1996;111(3):209-19.
19. de Groot M, Vernooij MW, Klein S, et al. Improving alignment in Tract-based spatial statistics: evaluation and optimization of image registration. Neuroimage. 2013;76:400-11.
20. Pastor-Bernier A 2022;Pages.

21. Horn A, Kuhn AA. Lead-DBS: a toolbox for deep brain stimulation electrode localizations and visualizations. Neuroimage. 2015;107:127-35.
22. Klapwijk ET, van de Kamp F, van der Meulen M, Peters S, Wierenga LM. Qoala-T: A supervised-learning tool for quality control of FreeSurfer segmented MRI data. Neuroimage. 2019;189:116-29.
23. Zhang Y, Larcher KM, Misic B, Dagher A. Anatomical and functional organization of the human substantia nigra and its connections. Elife. 2017;6.
24. Nowacki A, Barlatey S, Al-Fatly B, et al. Probabilistic Mapping Reveals Optimal Stimulation Site in Essential Tremor. Ann Neurol. 2022;91(5):602-12.
25. Neudorfer C, Kroneberg D, Al-Fatly B, et al. Personalizing Deep Brain Stimulation Using Advanced Imaging Sequences. Ann Neurol. 2022;91(5):613-28.
26. Horn A, Kuhn AA, Merkl A, Shih L, Alterman R, Fox M. Probabilistic conversion of neurosurgical DBS electrode coordinates into MNI space. Neuroimage. 2017;150:395-404.
27. Middlebrooks EH, Domingo RA, Vivas-Buitrago T, et al. Neuroimaging Advances in Deep Brain Stimulation: Review of Indications, Anatomy, and Brain Connectomics. AJNR Am J Neuroradiol. 2020;41(9):1558-68.
28. Horn A, Reich MM, Ewert S, et al. Optimal deep brain stimulation sites and networks for cervical vs. generalized dystonia. Proc Natl Acad Sci U S A. 2022;119(14):e2114985119.
29. Buckner RL, Krienen FM, Castellanos A, Diaz JC, Yeo BT. The organization of the human cerebellum estimated by intrinsic functional connectivity. J Neurophysiol. 2011;106(5):2322-45.
30. Neudorfer C, Germann J, Elias GJB, Gramer R, Boutet A, Lozano AM. A high-resolution in vivo magnetic resonance imaging atlas of the human hypothalamic region. Sci Data. 2020;7(1):305.
31. Boutet A, Germann J, Gwun D, et al. Sign-specific stimulation 'hot' and 'cold' spots in Parkinson's disease validated with machine learning. Brain Commun. 2021;3(2):fcab027.
32. Kumar VJ, van Oort E, Scheffler K, Beckmann CF, Grodd W. Functional anatomy of the human thalamus at rest. Neuroimage. 2017;147:678-91.
33. Elias GJB, Boutet A, Joel SE, et al. Probabilistic Mapping of Deep Brain Stimulation: Insights from 15 Years of Therapy. Ann Neurol. 2021;89(3):426-43.
34. Choi EY, Yeo BT, Buckner RL. The organization of the human striatum estimated by intrinsic functional connectivity. J Neurophysiol. 2012;108(8):2242-63.
35. Tang Y, Sun W, Toga AW, Ringman JM, Shi Y. A probabilistic atlas of human brainstem pathways based on connectome imaging data. Neuroimage. 2018;169:227-39.
36. Lau JC, Xiao Y, Haast RAM, et al. Direct visualization and characterization of the human zona incerta and surrounding structures. Hum Brain Mapp. 2020;41(16):4500-17.
37. Ewert S, Plettig P, Li N, et al. Toward defining deep brain stimulation targets in MNI space: A subcortical atlas based on multimodal MRI, histology and structural connectivity. Neuroimage. 2018;170:271-82.
38. Ilinsky I, Horn A, Paul-Gilloteaux P, Gressens P, Verney C, Kultas-Ilinsky K. Human Motor Thalamus Reconstructed in 3D from Continuous Sagittal Sections with Identified Subcortical Afferent Territories. eNeuro. 2018;5(3).
39. Tian Y, Margulies DS, Breakspear M, Zalesky A. Topographic organization of the human subcortex unveiled with functional connectivity gradients. Nat Neurosci. 2020;23(11):1421-32.

40. Su JH, Thomas FT, Kasoff WS, et al. Thalamus Optimized Multi Atlas Segmentation (THOMAS): fast, fully automated segmentation of thalamic nuclei from structural MRI. Neuroimage. 2019;194:272-82.
41. He X, Chaitanya G, Asma B, et al. Disrupted basal ganglia-thalamocortical loops in focal to bilateral tonic-clonic seizures. Brain. 2020;143(1):175-90.
42. Alkemade A, Mulder MJ, Groot JM, et al. The Amsterdam Ultra-high field adult lifespan database (AHEAD): A freely available multimodal 7 Tesla submillimeter magnetic resonance imaging database. Neuroimage. 2020;221:117200.
43. Yu B, Li L, Guan X, et al. HybraPD atlas: Towards precise subcortical nuclei segmentation using multimodality medical images in patients with Parkinson disease. Hum Brain Mapp. 2021;42(13):4399-421.
44. Xiao Y, Lau JC, Anderson T, et al. An accurate registration of the BigBrain dataset with the MNI PD25 and ICBM152 atlases. Sci Data. 2019;6(1):210.
45. Furtjes AE, Cole JH, Couvy-Duchesne B, Ritchie SJ. A quantified comparison of cortical atlases on the basis of trait morphometricity. Cortex. 2023;158:110-26.
46. Alfaro-Almagro F, McCarthy P, Afyouni S, et al. Confound modelling in UK Biobank brain imaging. Neuroimage. 2021;224:117002.
47. Collister JA, Liu X, Clifton L. Calculating Polygenic Risk Scores (PRS) in UK Biobank: A Practical Guide for Epidemiologists. Front Genet. 2022;13:818574.
