## supplemental assessment of environmental risk for "Brain imaging phenotypes associated with polygenic risk for Essential Tremor"

In our analysis of high-risk healthy controls and low-risk healthy controls compared to ET patients, we suggest that while the high-risk controls do carry increased genetic risk towards ET, they do not pass the threshold for disease and thus do not display tremors. This likely means that these individuals have not yet accumulated a sufficient combination of both genetic and environmental risk to push them past that disease threshold. The PRS largely captured the genetic component of ET risk but could not capture any environmental risk factors. Therefore, we decided to compare different lifestyle factor reports taken from the UKB to look for differences in environmental factors between the high-risk controls, low-risk controls, and ET patients. The lifestyle factor responses used came from the first imaging visit for UKB participants. The lifestyle factors chosen for study were previously outlined by Ong *et. al.*[1]: alcohol intake, smoking behavior, caffeine consumption, antioxidants (vitamin E and C consumption used as a proxy), beta-carboline alkaloids (meat consumption and caffeine used as a proxy), and pesticide exposures.

No significant differences between cohorts were observed. Meat consumption appeared to approach significance (Fisher's exact test: p-value: 0.053), but this is likely due to the very small number of reports in the low-risk control group which may have biased results. There did not appear to be a difference between the high-risk group and ET patients. Interestingly, vitamin C levels between cases and high-risk individuals neared significance (Wilcoxon rank sum test: p-value: 0.077) but did not meet the cut-off. However, sample sizes were small for many analyses due to incomplete survey responses, thus no meaningful conclusions on differing environmental exposures can be drawn. Due to this limitation these analyses are speculative. Future studies examining the differences between environmental factors between high genetic risk controls and cases may shed light on which environmental factors are more critical in causing patients to develop ET.

**<u>Lifestyle Factor Tests Table of Contents:</u>**

*Alcohol Intake Frequency*........................................*p.2*

*Smoking Status*........................................*p.3*

*Caffeine Consumption*........................................*p.4*

*Meat Consumption*........................................*p.5*

*Vitamin E Intake*........................................*p.6*

*Vitamin E Intake (Women)*........................................*p.7*

*Vitamin E Intake (Men)*........................................*p.8*

*Vitamin C Intake*........................................*p.9*

*Vitamin C Intake (Women)*........................................*p.10*

*Vitamin C Intake (Men)*........................................*p.11*

*(Note: No cases or controls reported being exposed to pesticides.)*

**Lifestyle factors/environmental exposure tests:**

*Phenotype: Alcohol Intake Frequency*

UKB data-field: 1558 (Self report. Touchscreen question: "About how often do you drink alcohol?" If the participant activated the Help button they were shown the message: If this varies a lot, please provide an average considering your intake over the last year)

Data-field responses by cohort phenotype:

| Cohort Response | ET Cohort | High-Risk Cohort | Low-Risk Cohort |
|---|---|---|---|
| Daily or almost daily | 13 | 10 | 14 |
| Three or four times a week | 10 | 15 | 13 |
| Once or twice a week | 7 | 11 | 9 |
| One to three times a month | 8 | 8 | 5 |
| Special occasions only | 6 | 4 | 4 |
| Never | 5 | 1 | 3 |
| N/A | 0 | 0 | 1 |

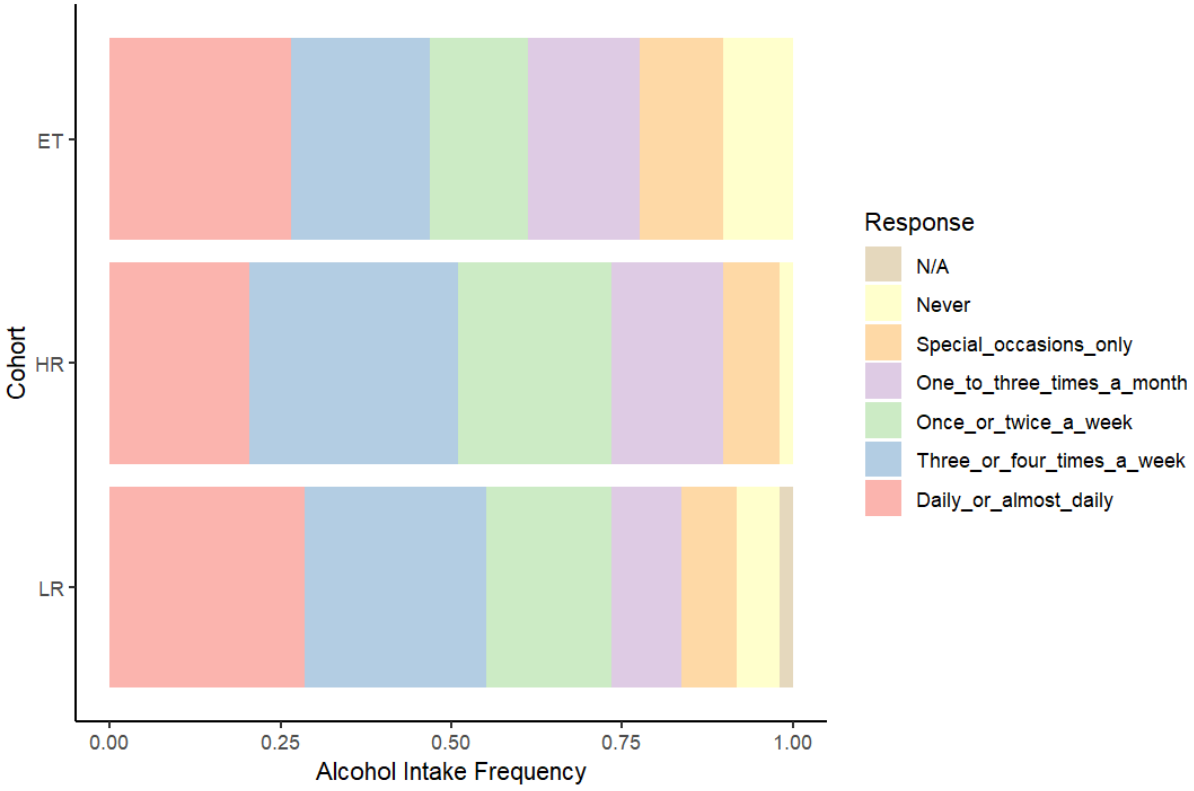


**Statistical test**: Fisher's Exact Test for Count Data
**p-value**: 0.7691

*Phenotype: Smoking Status*

UKB data-field: 20116 (Self report. Touchscreen questionnaire. This field summarises the current/past smoking status of the participant.)

Data-field responses by cohort phenotype:

| Cohort Response | ET Cohort | High-Risk Cohort | Low-Risk Cohort |
|---|---|---|---|
| Never | 34 | 35 | 29 |
| Previous | 12 | 13 | 18 |
| Current | 3 | 1 | 1 |
| N/A | 0 | 0 | 1 |

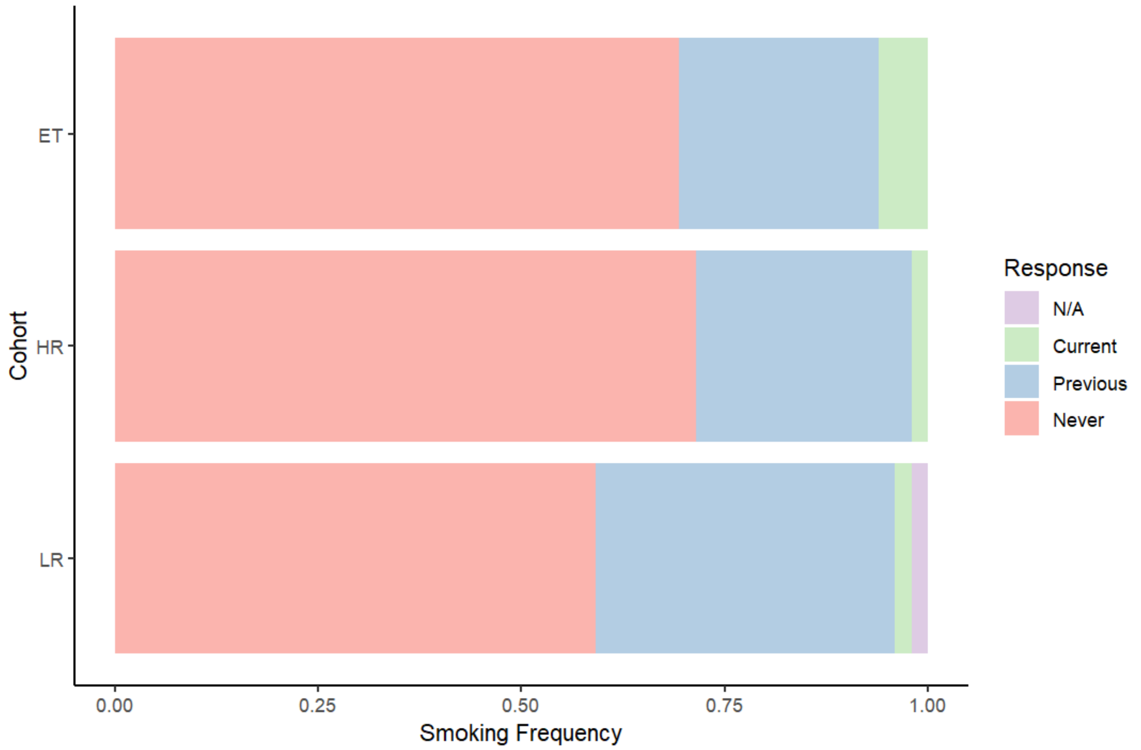


**Statistical test**: Fisher's Exact Test for Count Data
**p-value**: 0.5177

*Phenotype: Caffeine Consumption within Last Hour*

UKB data-field: 3089 (Self report. Participants were asked whether they had drank caffeine within the hour prior to doing a spirometry test.)

Data-field responses by cohort phenotype:

| Cohort Response | ET Cohort | High-Risk Cohort | Low-Risk Cohort |
|---|---|---|---|
| Yes | 9 | 11 | 11 |
| No | 32 | 35 | 34 |
| N/A | 8 | 3 | 4 |

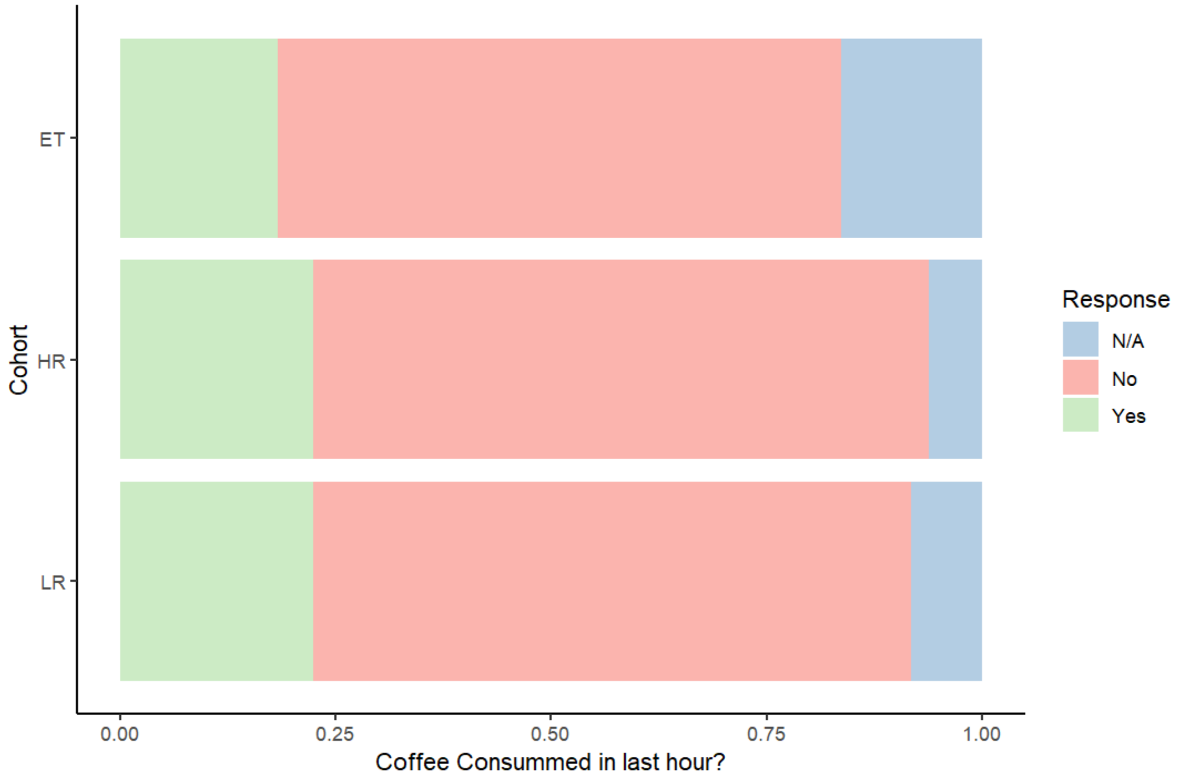


**Statistical test**: Fisher's Exact Test for Count Data
**p-value**: 0.9669

*Phenotype: Meat Consumption*

UKB data-field: 103000 (Self report. Question asked: "Did you eat any meat or poultry yesterday? Think about curry, stir-fry, sandwiches, pie fillings, sausages/burgers, liver, pate or mince." If the participant activated the Help feature they were shown the message: *Select which category best describes your type of meat or poultry, otherwise select the Other category. Game and offal (not liver) should be recorded in the Other category.*)

Data-field responses by cohort phenotype:

| Cohort Response | ET Cohort | High-Risk Cohort | Low-Risk Cohort |
|---|---|---|---|
| Yes | 16 | 18 | 6 |
| No | 3 | 5 | 7 |
| N/A | 30 | 26 | 36 |

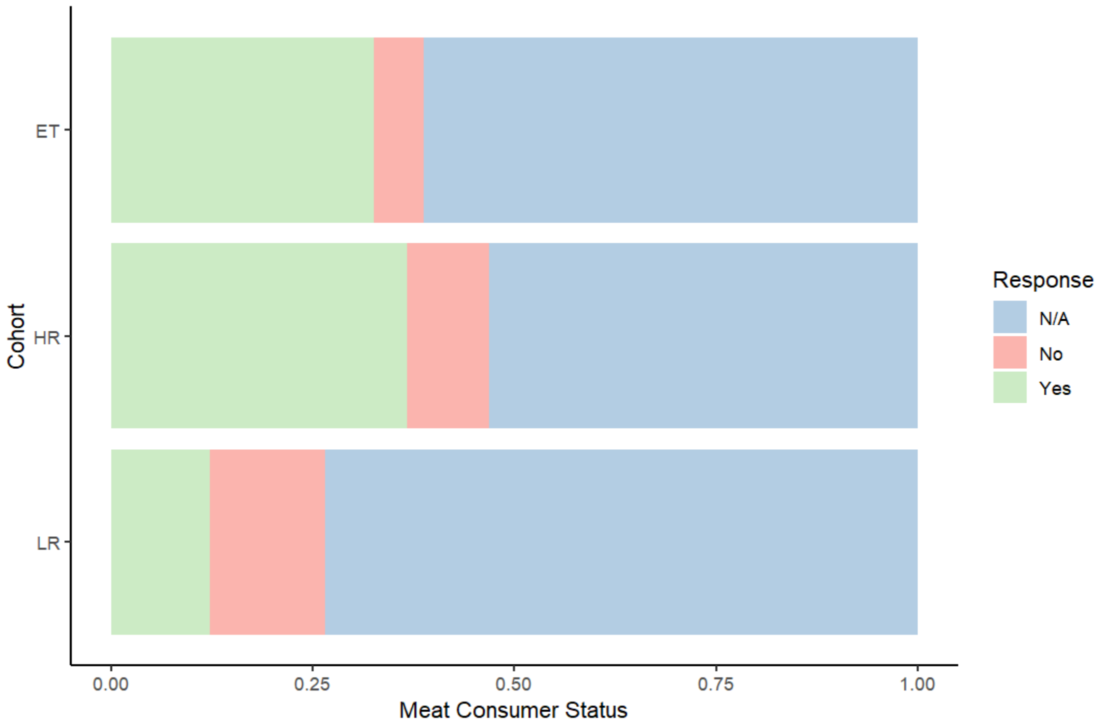


**Statistical test**: Fisher's Exact Test for Count Data
**p-value**: 0.05316

*Phenotype: Vitamin E*

UKB data-field: 26028 (Self report. Vitamin E from overall diet. Estimated intake from past 24 hours)

Data-field responses by cohort phenotype:

| Cohort Response | ET Cohort (n=19) | High-Risk Cohort (n=23) | Low-Risk Cohort (n=13) |
|---|---|---|---|
| Minimum | 1.870 | 3.200 | 0.28 |
| 1st Quantile | 5.450 | 6.090 | 5.28 |
| Median | 7.850 | 9.060 | 9.05 |
| Mean | 8.894 | 9.524 | 10.30 |
| 3rd Quantile | 9.460 | 12.485 | 12.35 |
| Max | 38.110 | 17.770 | 29.95 |

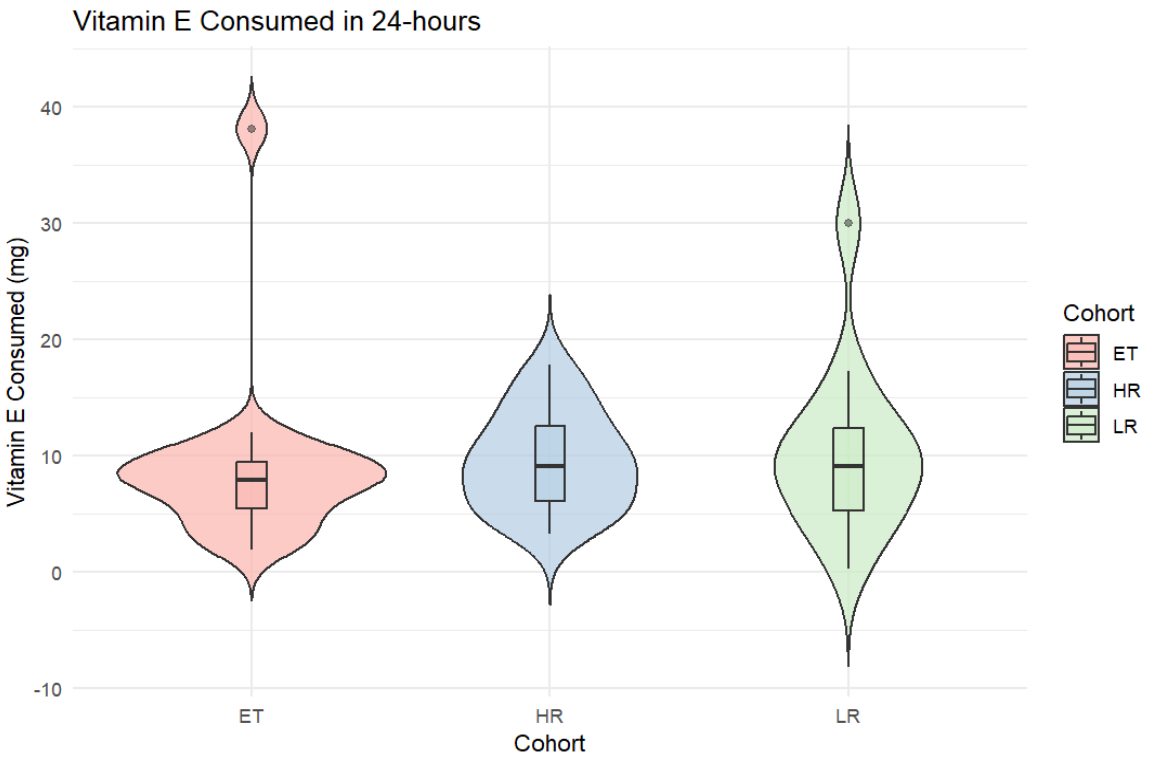


**Statistical test**: Kruskal-Wallis rank sum test
Kruskal-Wallis chi-squared = 1.9142, df = 2,
**p-value**: 0.384

Because dietary requirements may vary between men and women, we re-did the analysis in the subdivision of men and women.

*Vitamin E intake in **Women**:*

Data-field responses by cohort phenotype:

| Cohort Response | ET Cohort (n=9) | High-Risk Cohort (n=8) | Low-Risk Cohort (n=6) |
|---|---|---|---|
| Minimum | 1.87 | 3.970 | 3.650 |
| 1st Quantile | 6.08 | 6.338 | 6.030 |
| Median | 7.58 | 9.855 | 8.280 |
| Mean | 10.21 | 9.453 | 7.568 |
| 3rd Quantile | 8.87 | 11.260 | 9.300 |
| Max | 38.11 | 16.810 | 10.280 |

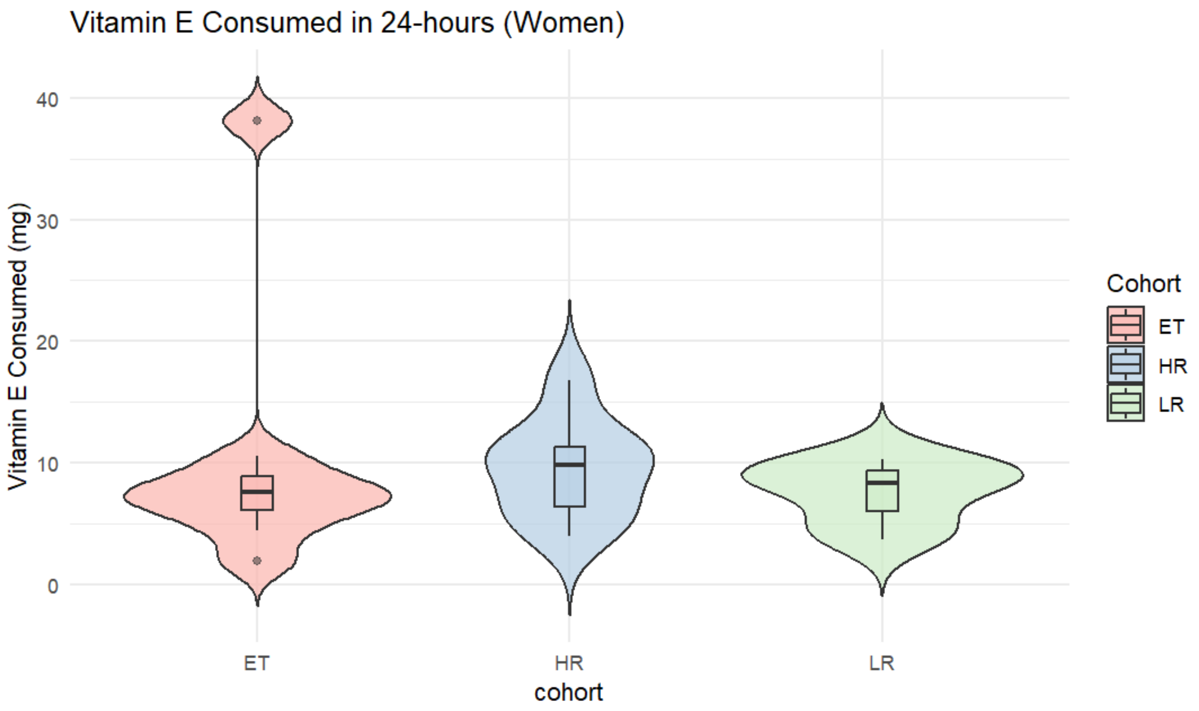


**Statistical test**: Kruskal-Wallis rank sum test
Kruskal-Wallis chi-squared = 1.3509, df = 2,
**p-value**: 0.5089

We then compared just the ET cohort to the high-risk (HR) cohort:

**Statistical test**: Wilcoxon rank sum exact test
W = 27,
**p-value**: 0.4234

*Vitamin E intake in **Men**:*

Data-field responses by cohort phenotype:

| Cohort Response | ET Cohort (n=10) | High-Risk Cohort (n=15) | Low-Risk Cohort (n=7) |
|---|---|---|---|
| Minimum | 2.800 | 3.200 | 4.89 |
| 1st Quantile | 5.577 | 5.900 | 10.30 |
| Median | 8.585 | 8.580 | 12.35 |
| Mean | 7.711 | 9.562 | 14.08 |
| 3rd Quantile | 9.715 | 13.250 | 15.37 |
| Max | 11.950 | 17.770 | 29.95 |

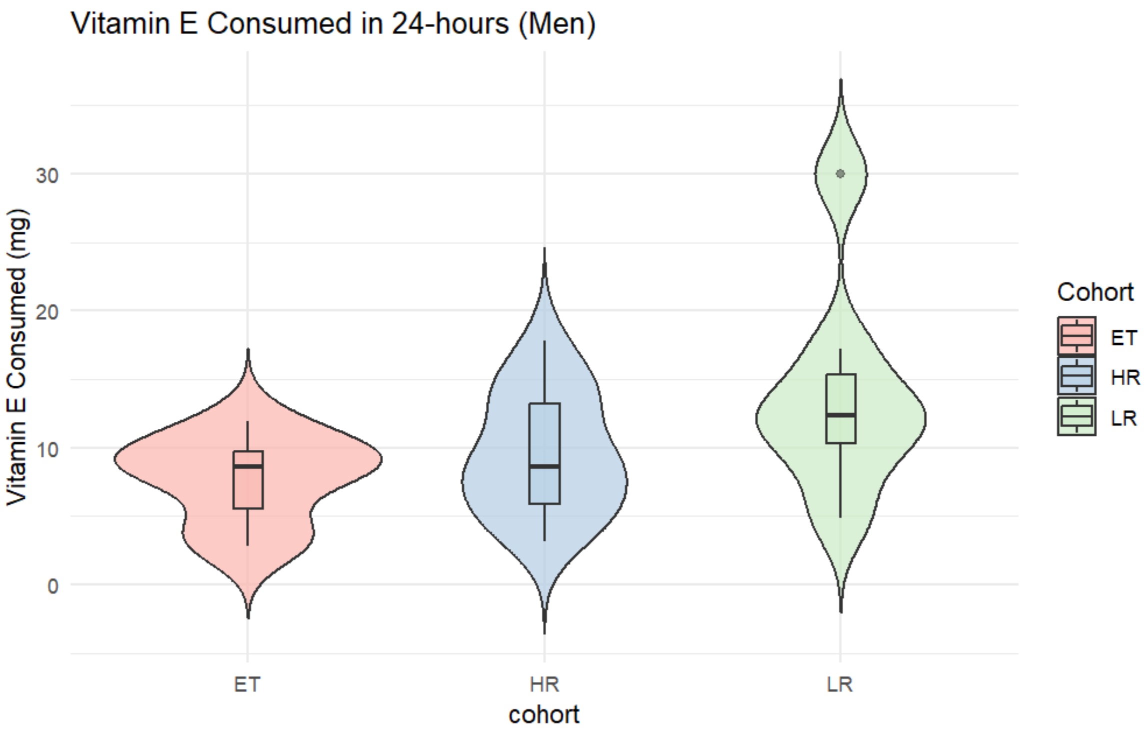


**Statistical test**: Kruskal-Wallis rank sum test
Kruskal-Wallis chi-squared = 4.7551, df = 2,
**p-value**: 0.0928

We then compared just the ET cohort to the high-risk (HR) cohort:

**Statistical test**: Wilcoxon rank sum exact test
W = 59,
**p-value**: 0.3969

*Phenotype: Vitamin C*

UKB data-field: 26023 (Self report. Vitamin E from overall diet. Estimated intake from past 24 hours)

Data-field responses by cohort phenotype:

| Cohort Response | ET Cohort (n=19) | High-Risk Cohort (n=23) | Low-Risk Cohort (n=13) |
|---|---|---|---|
| Minimum | 9.95 | 28.26 | 32.57 |
| 1st Quantile | 44.20 | 97.55 | 94.59 |
| Median | 78.40 | 147.45 | 148.66 |
| Mean | 116.45 | 163.74 | 143.20 |
| 3rd Quantile | 154.88 | 226.87 | 209.02 |
| Max | 371.45 | 344.54 | 276.25 |

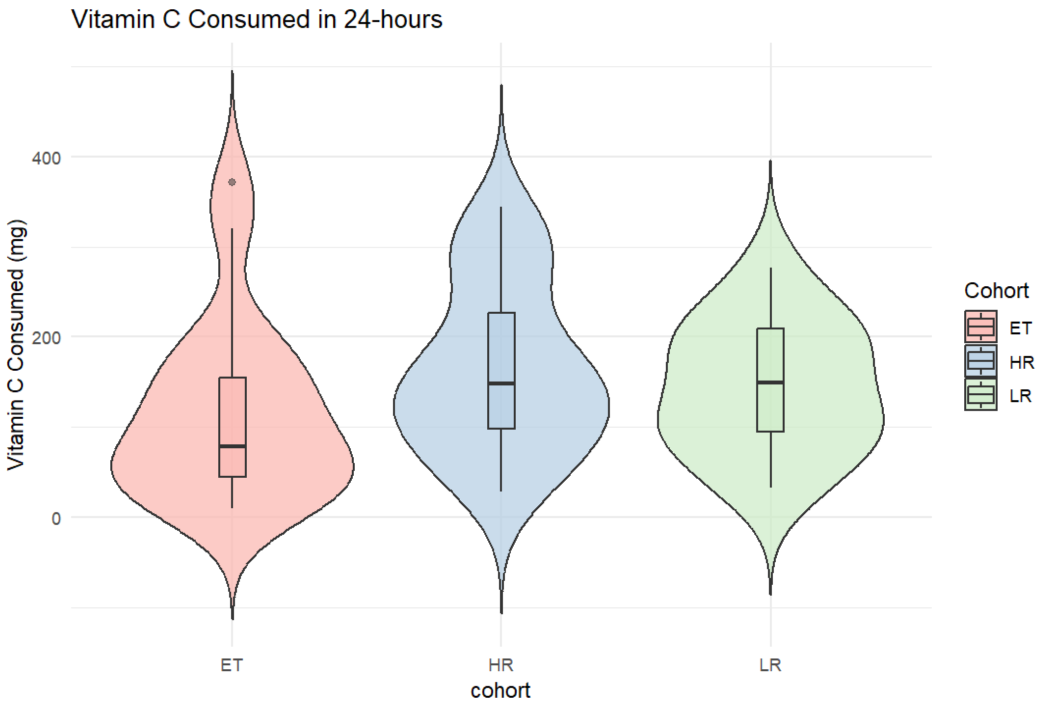


**Statistical test**: Kruskal-Wallis rank sum test
Kruskal-Wallis chi-squared = 3.6295, df = 2,
**p-value**: 0.1629

We then compared just the ET cohort to the high-risk (HR) cohort:

**Statistical test**: Wilcoxon rank sum exact test
W = 148,
**p-value**: 0.07676

Because dietary requirements may vary between men and women, we re-did the analysis in the subdivision of men and women.

*Vitamin C intake in **Women***:

Data-field responses by cohort phenotype:

| Cohort Response | ET Cohort (n=9) | High-Risk Cohort (n=8) | Low-Risk Cohort (n=6) |
|---|---|---|---|
| Minimum | 9.95 | 28.26 | 32.57 |
| 1st Quantile | 23.30 | 104.07 | 99.59 |
| Median | 78.22 | 170.91 | 129.62 |
| Mean | 135.67 | 179.19 | 130.32 |
| 3rd Quantile | 196.60 | 265.24 | 171.04 |
| Max | 371.45 | 308.00 | 216.71 |

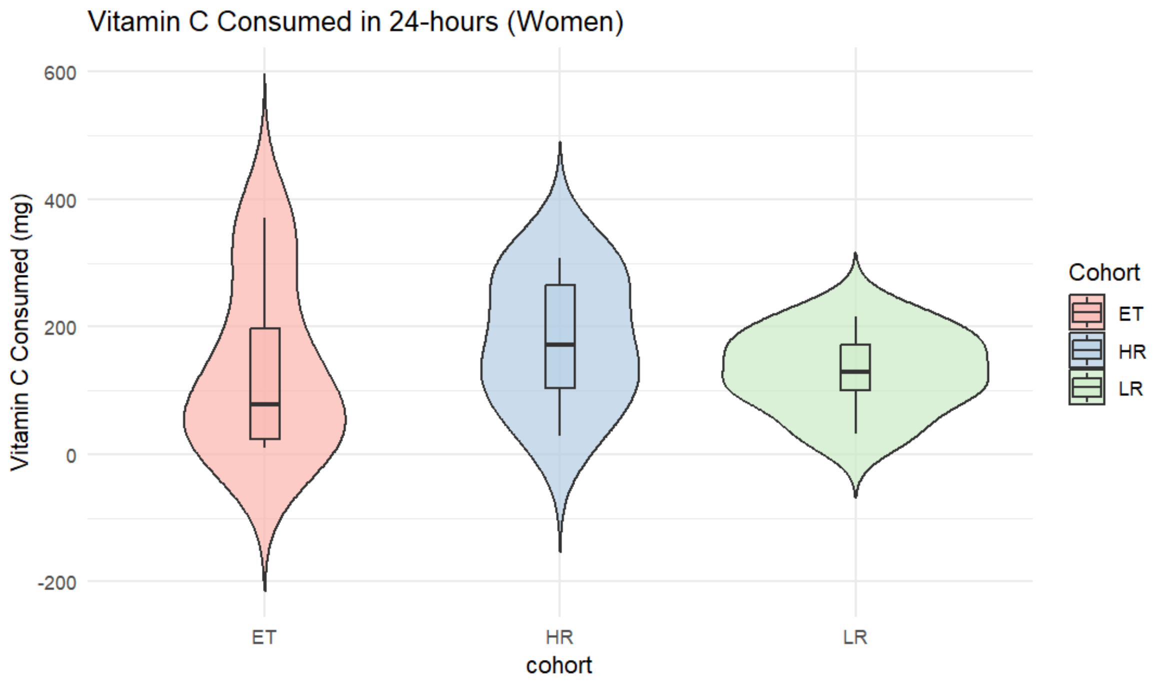


**Statistical test**: Kruskal-Wallis rank sum test
Kruskal-Wallis chi-squared = 1.2428, df = 2,
**p-value**: 0.5372

We then compared just the ET cohort to the high-risk (HR) cohort:

**Statistical test**: Wilcoxon rank sum exact test
W = 26,
**p-value**: 0.3704

*Vitamin C intake in **Men**:*

Data-field responses by cohort phenotype:

| Cohort Response | ET Cohort (n=10) | High-Risk Cohort (n=15) | Low-Risk Cohort (n=7) |
|---|---|---|---|
| Minimum | 20.93 | 33.22 | 44.32 |
| 1st Quantile | 56.78 | 93.25 | 92.23 |
| Median | 94.56 | 131.37 | 148.66 |
| Mean | 99.15 | 155.50 | 154.23 |
| 3rd Quantile | 144.03 | 187.90 | 212.96 |
| Max | 196.92 | 344.54 | 276.25 |

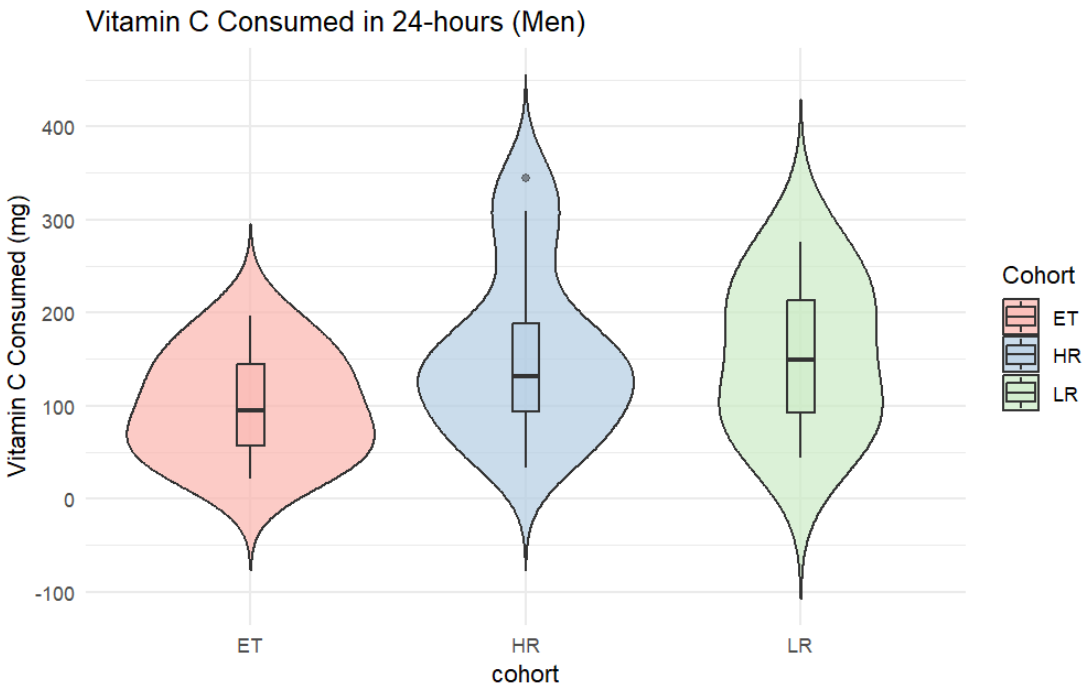


**Statistical test**: Kruskal-Wallis rank sum test
Kruskal-Wallis chi-squared = 2.9391, df = 2,
**p-value**: 0.23

We then compared just the ET cohort to the high-risk (HR) cohort:

**Statistical test**: Wilcoxon rank sum exact test
W = 47,
**p-value**: 0.1289

We used the threshold liability model to contextualize ET being a complex disorder though a theoretical normal distribution of risk towards ET, which we can call the liability distribution. There exists a certain point in this liability distribution, known as the disease threshold, that can separate healthy individuals from affected individuals. Anyone below this threshold is unaffected, while anyone above is affected with the condition. Within this framework, both genetic risk and environmental risk increases one's liability and can bring one closer to the disease threshold. One reason that some people may not express tremor but still carry high genetic risk could be that some environmental risk is needed to take those individuals pass that threshold to develop tremors. The lifestyle factors we tested however, do not provide us with answer of which of the established environmental risk and protective factors (alcohol intake, smoking behavior, caffeine consumption, antioxidants, beta-carboline alkaloids, and pesticide exposures) are responsible for lack of tremors in the high-risk control group compares to ET cases. Higher powered future studies are needed to further investigate this.

Still, to explain the lack of tremors in the high-risk cohort compared to ET cases, we can consider the challenge from another lens. Viewing things from a more biological angle, we can think that even though these healthy individuals carry a higher genetic risk towards ET, perhaps the combination of risk alleles that they carry are not complementary. For instance, if an ET patient carries X number of risk variants but they're mostly localized to genes in just a few pathways, that patient may be more affected than a healthy individual who has the same number of X risk variants but happen to be diluted across many different pathways. Future studies could use a larger cohort of whole genome sequenced patients and high-risk controls to identify variants for enrichment assays to see how greater burden of variants in a given pathway could affect disease manifestation. This would also enable the discovery of potential rare variants with a greater burden in the ET cohort than controls which may explain the onset of tremors.

Interestingly, previous studies found that first-degree relatives of ET patients exhibited greater tremor scores than individuals with no affected relatives, suggesting that a subclinical ET may exist where penetrance is still not complete.[2,3] A study examining the burden of high-risk and low-risk unaffected individuals (informed by ET family history), high-risk individuals were up to 4.5 more likely to exhibit a mild sub-clinical tremor compared to their low-risk counterparts.[2] The authors suggested that the high burden of mild tremor in the high-risk group may be a *forme fruste* of ET.[2] It is possible as the high-risk controls in our study do possess a greater risk towards ET and share similar cerebellar abnormalities, that they may possess a subclinical version of ET perhaps even with mild tremors. These high-risk individuals may benefit from reduced penetrance or variable expressivity of risk variants, which ultimately contributes to a degree of grey matter vulnerability insufficient the development of ET unless additional risk factors are accumulated.

### References:


1. Ong YL, Deng X, Tan EK**.** Etiologic links between environmental and lifestyle factors and Essential tremor. Ann Clin Transl Neurol. 2019;6(5):979-89.
2. Louis ED, Meyers JH, Badejo OM, Cristal AD, Hickman R, Factor-Litvak P**.** Comparative Burden of Subclinical Tremor in a Cohort of Normal Individuals Stratified by Familial Risk for Essential Tremor. Neuroepidemiology. 2018;50(1-2):41-6.
3. Louis ED, Ford B, Frucht S, Ottman R**.** Mild tremor in relatives of patients with essential tremor: what does this tell us about the penetrance of the disease? Arch Neurol. 2001;58(10):1584-9.
