## Supplementary Figures for "Brain imaging phenotypes associated with polygenic risk for Essential Tremor"

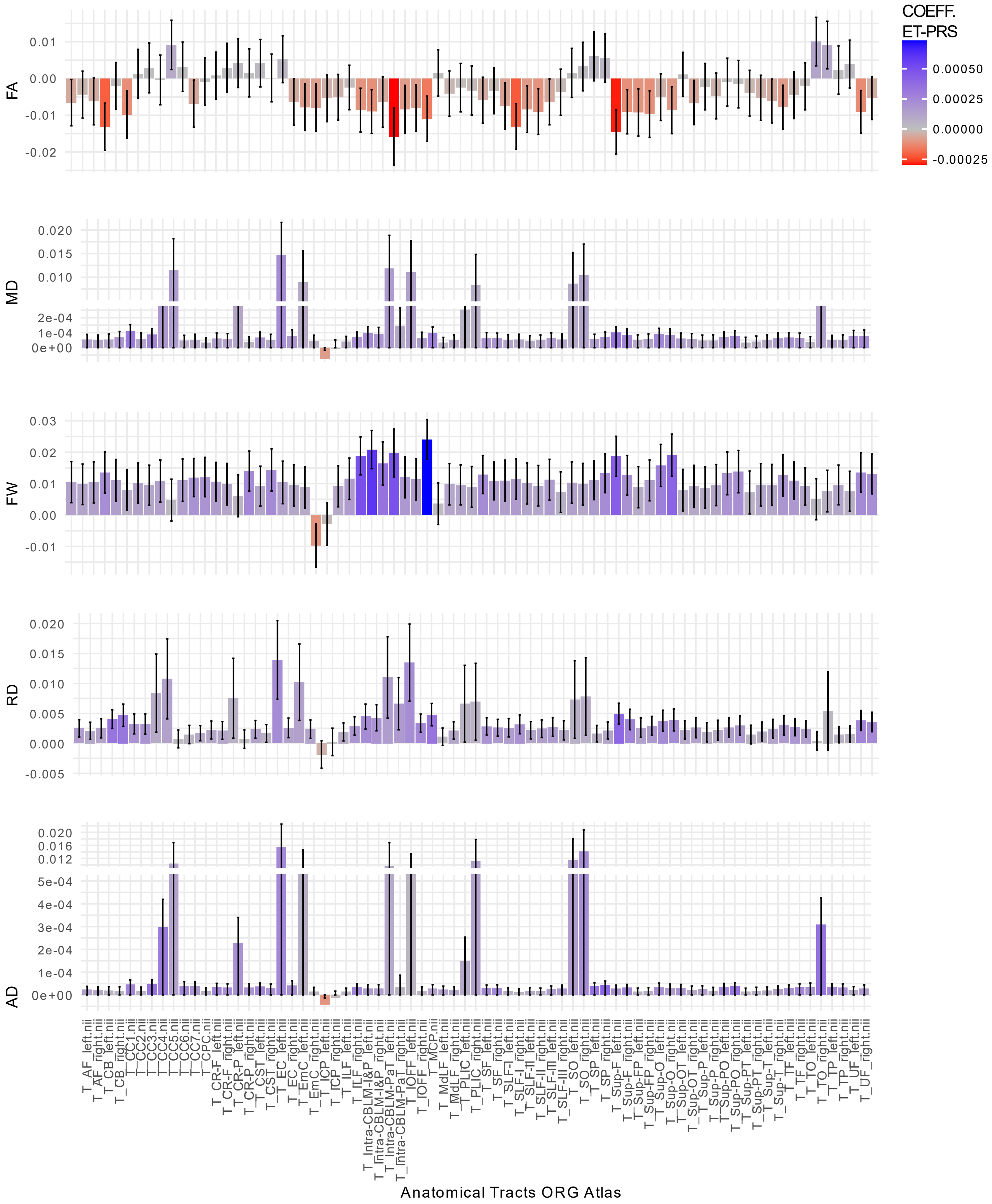


**Figure S1. Histograms ET PRS associations with white matter diffusion-weighted magnetic resonance imaging (ORG Atlas)**

Histograms representing the associations of ET polygenic risk scores (PRS) (coefficients) with white matter microstructure measures in 73 anatomical tracts (ORG atlas). Measures of fractional anisotropy (FA), mean diffusivity (MD), free water (FW), radial diffusivity (RD), and axial diffusivity (AD).

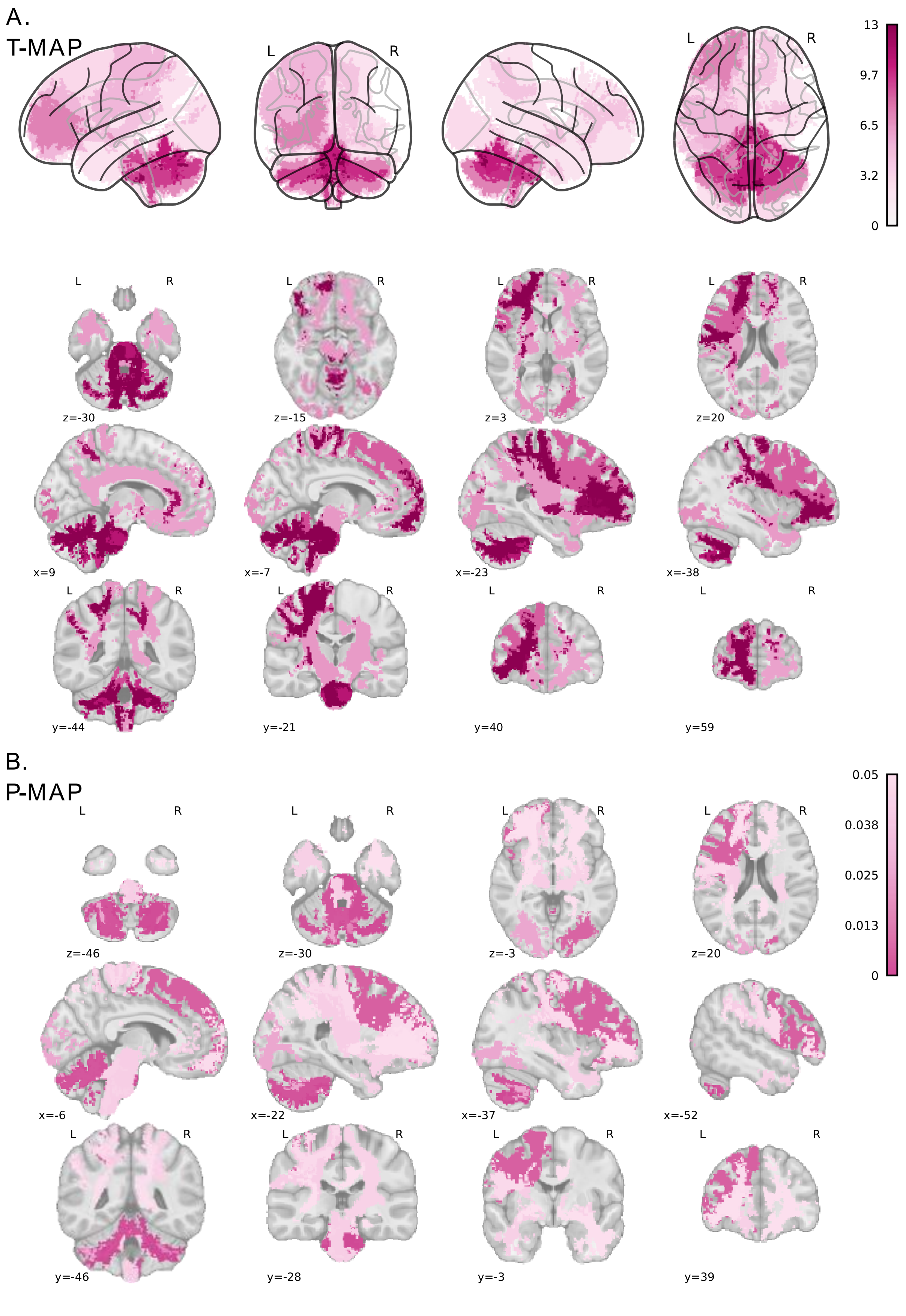


**Figure S2. White matter diffusion-weighted magnetic resonance imaging of free water in ORG Atlas**
Free water (FW) across white matter anatomical tracts reveals positive associations with ET polygenic risk scores (PRS) in cerebellar spinal and frontal projection tracts (ORG anatomically curated fiber clustering atlas, Zhang et al., 2018). (A) Glassbrain representations of the t-statistic projections indicate stronger positive associations in the cerebellum. ORG altas representation of the t-statistic in the MNI 152_nlin_asym_09 template and axial, transverse and coronal slices is shown below. (B) Mosaic representation of the FDR-adjusted p-value projections indicate positive associations in the spinal, cerebellar, and bilateral frontal lobe projections.

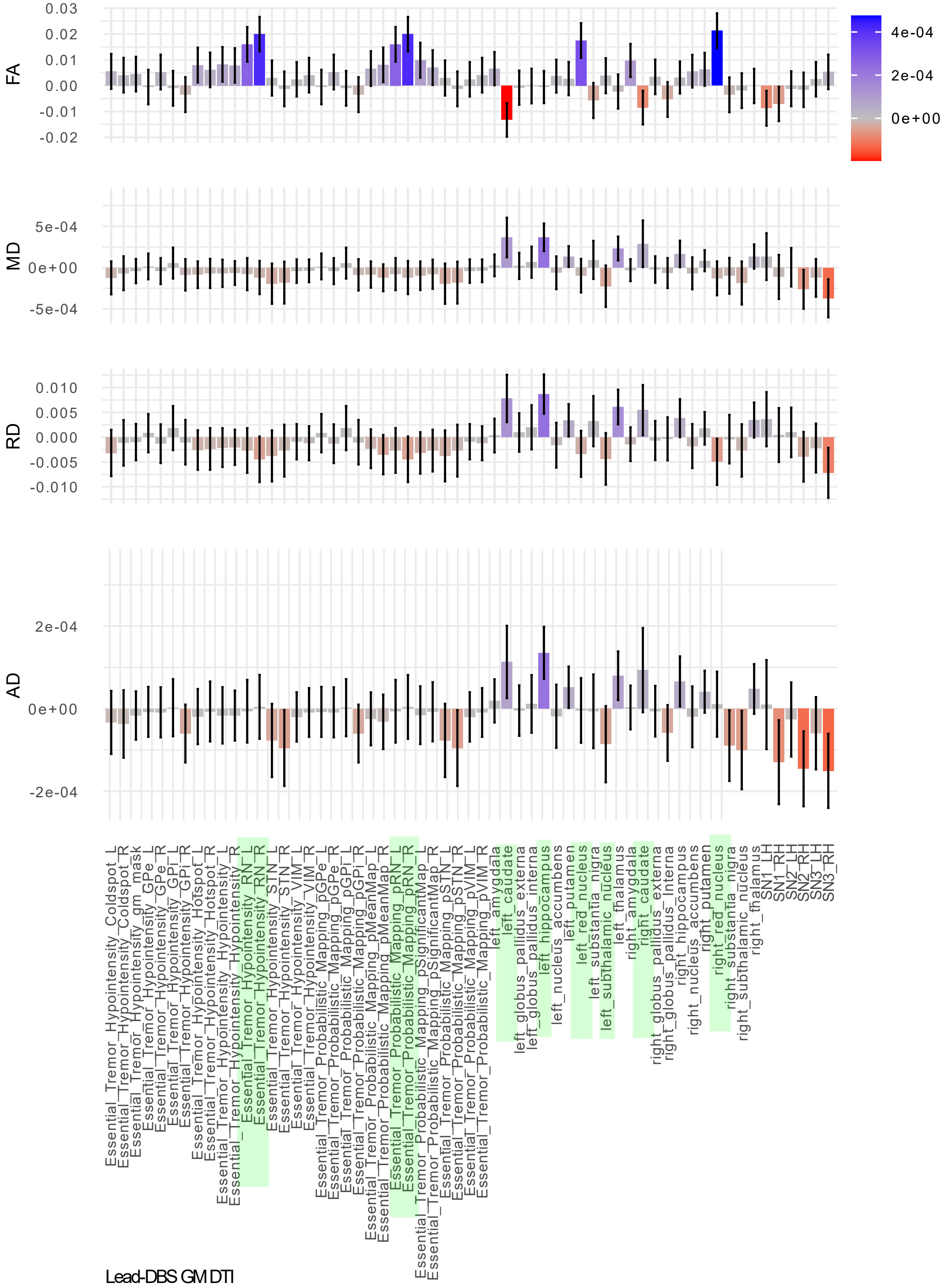


**Figure S3. Histograms ET PRS associations with grey matter diffusion-weighted magnetic resonance imaging in LeadDBS – red nucleus**

ET polygenic risk scores (PRS) regression coefficient histograms across Lead_DBS atlases reveal red nucleus associations in fractional anisotropy (FA) and thalamic, striatal associations in mean diffusivity (MD). Measures of fractional anisotropy (FA), mean diffusivity (MD), radial diffusivity (RD), and axial diffusivity (AD).

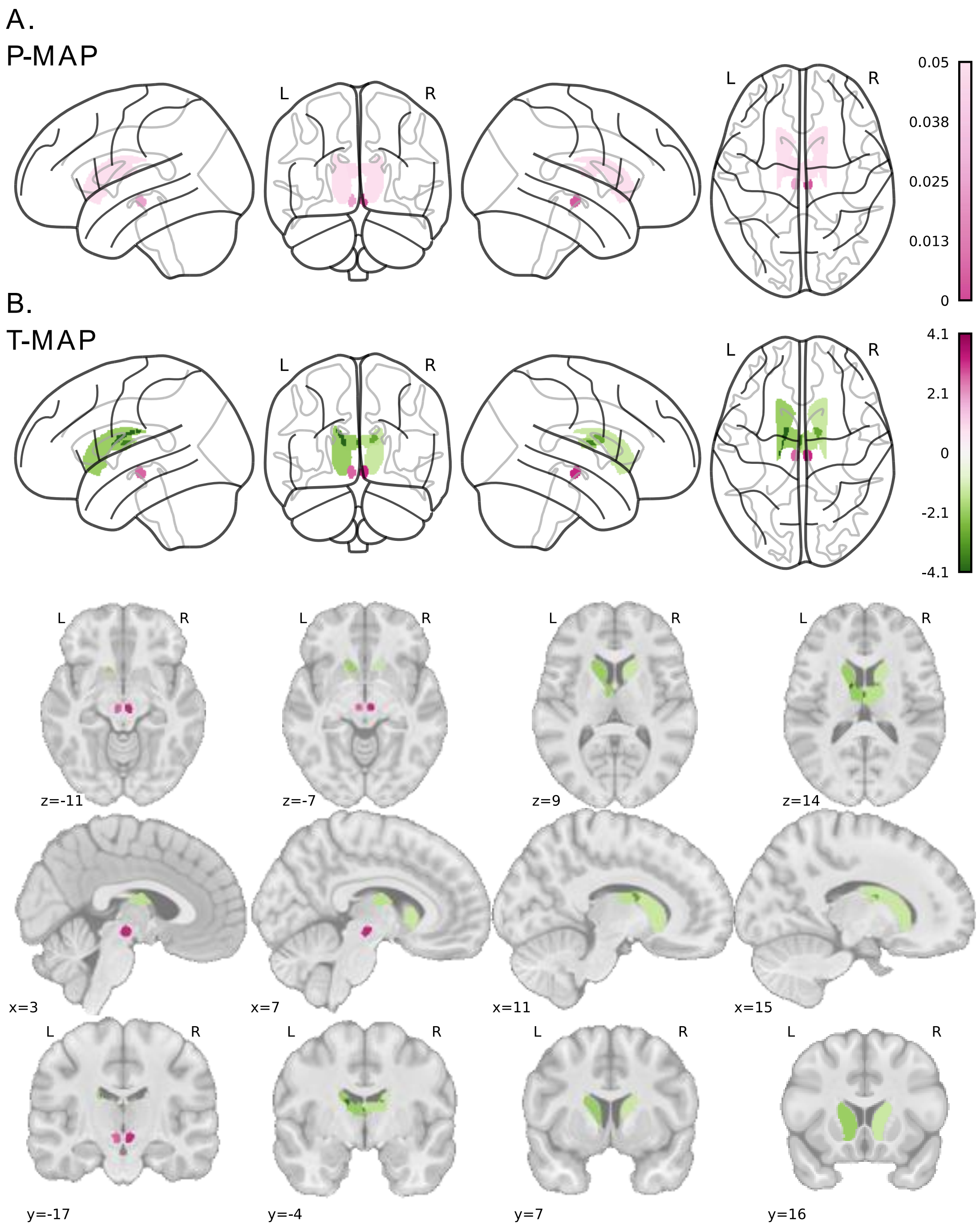


**Figure S4. Grey matter fractional anisotropy for red nucleus**

Fractional anisotropy (FA) in red nucleus (RN) and caudate (Lead-DBS Essential Tremor Probabilistic Mapping Nowacki 2022, and Essential Tremor Hypointensity, Neudorfer 2022) showing significant associations with ET polygenic risk score (PRS). (A) Glassbrain representation of the FDR adjusted p-value projections indicate stronger associations of FA and ET PRS in the bilateral RN. (B) Glassbrain representations of the t-statistic projections in these two regions indicate a positive association of FA in the RN and a negative association in the caudate. Mosaic representation of the t-statistic in the MNI I 152_nlin_asym_09c template and axial, transverse and coronal slices is shown below.

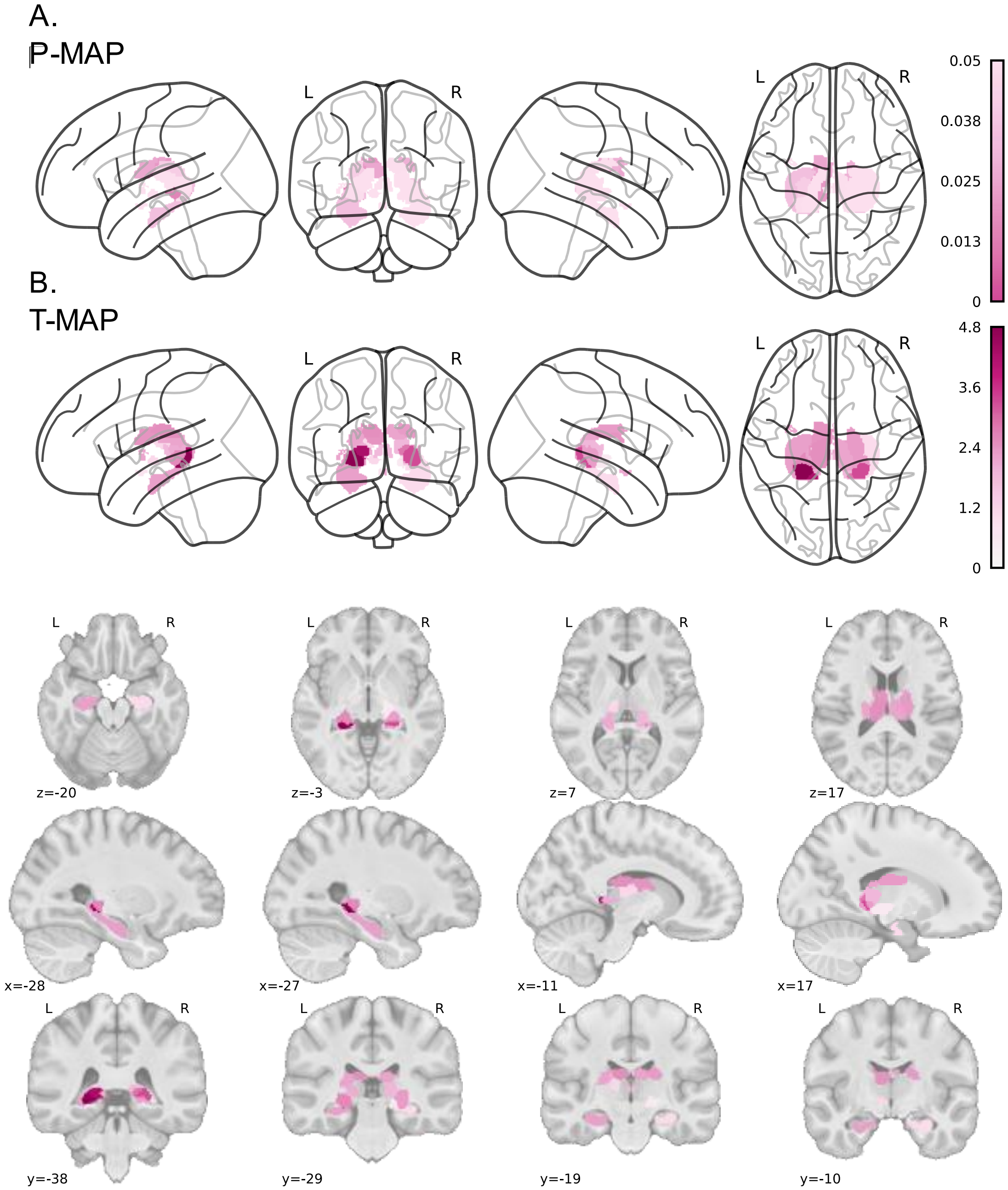


**Figure S5. Grey matter mean diffusivity for thalamus and hippocampus**

Mean diffusivity (MD) in functional thalamus and hippocampus (Lead-DBS Atlas of the Human Hypothalamus, Neudorfer & Germann 2020; Thalamic Functional Atlas Kumar 2017) showing significant associations with ET polygenic risk scores (PRS). (A) Glassbrain representation of the FDR adjusted p-value projections indicate stronger positive associations in the thalamus and weaker hypothalamic and hippocampus. (B) Glassbrain representations of the t-statistic projections indicate stronger positive associations in the posterior and ventral thalamic nuclei. A mosaic representation of the t-statistic in the MNI I 152_nlin_asym_09 template and axial, transverse and coronal slices is shown below. Fig. 5A. ET PRS shows negative associations with cortical volume in superior parietal and superior frontal areas, in addition to precuneus and posterior cingulate areas. Freesurfer DK parcellation (Desikan et al., 2006).

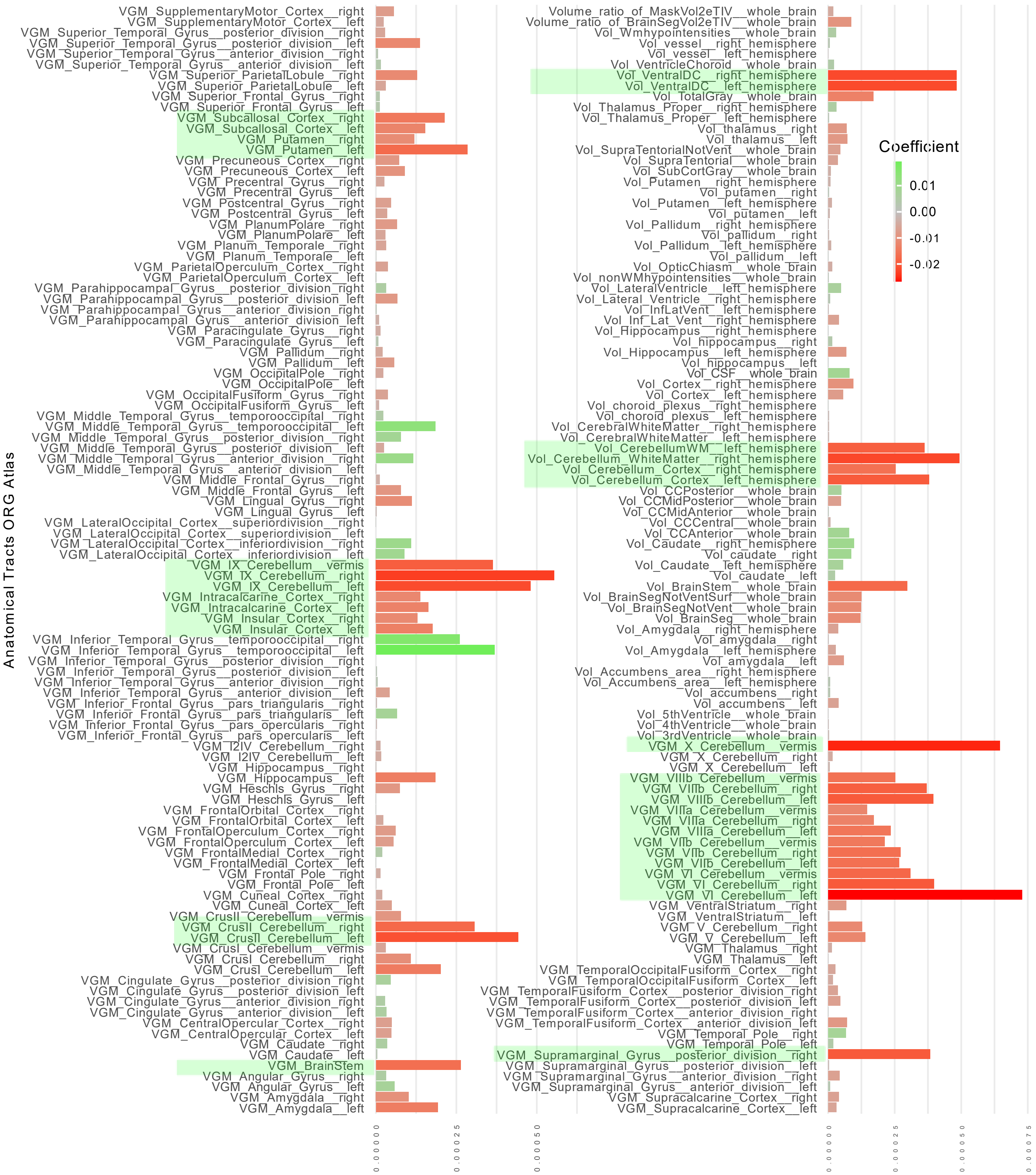


**Figure S6. Grey matter fractional anisotropy bar plots for red nucleus**

Fractional anisotropy (FA) in red nucleus (RN) and caudate (Lead-DBS Essential Tremor Probabilistic Mapping Nowacki 2022, and Essential Tremor Hypointensity, Neudorfer 2022) showing significant associations with ET polygenic risk scores (PRS).

A

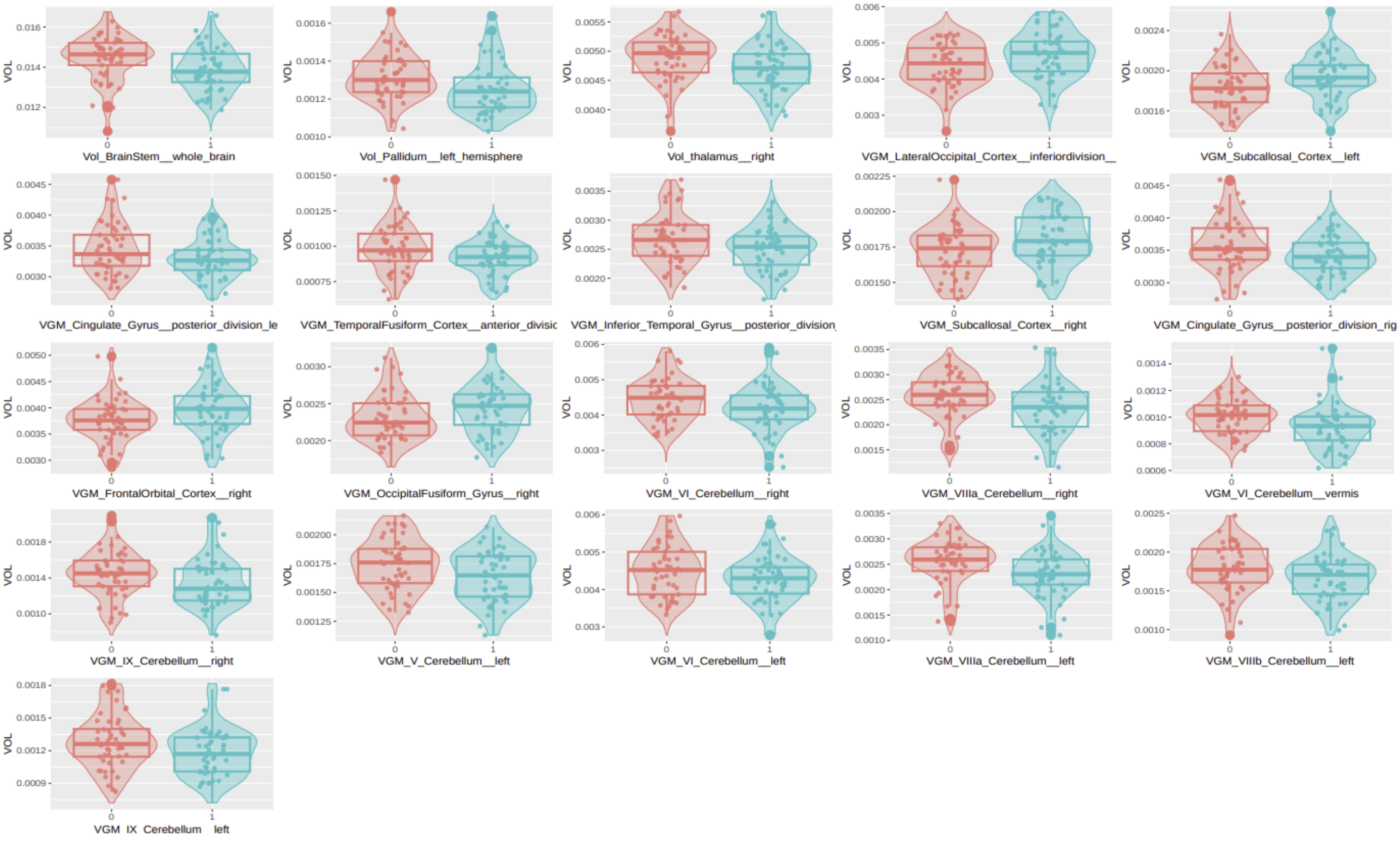


B

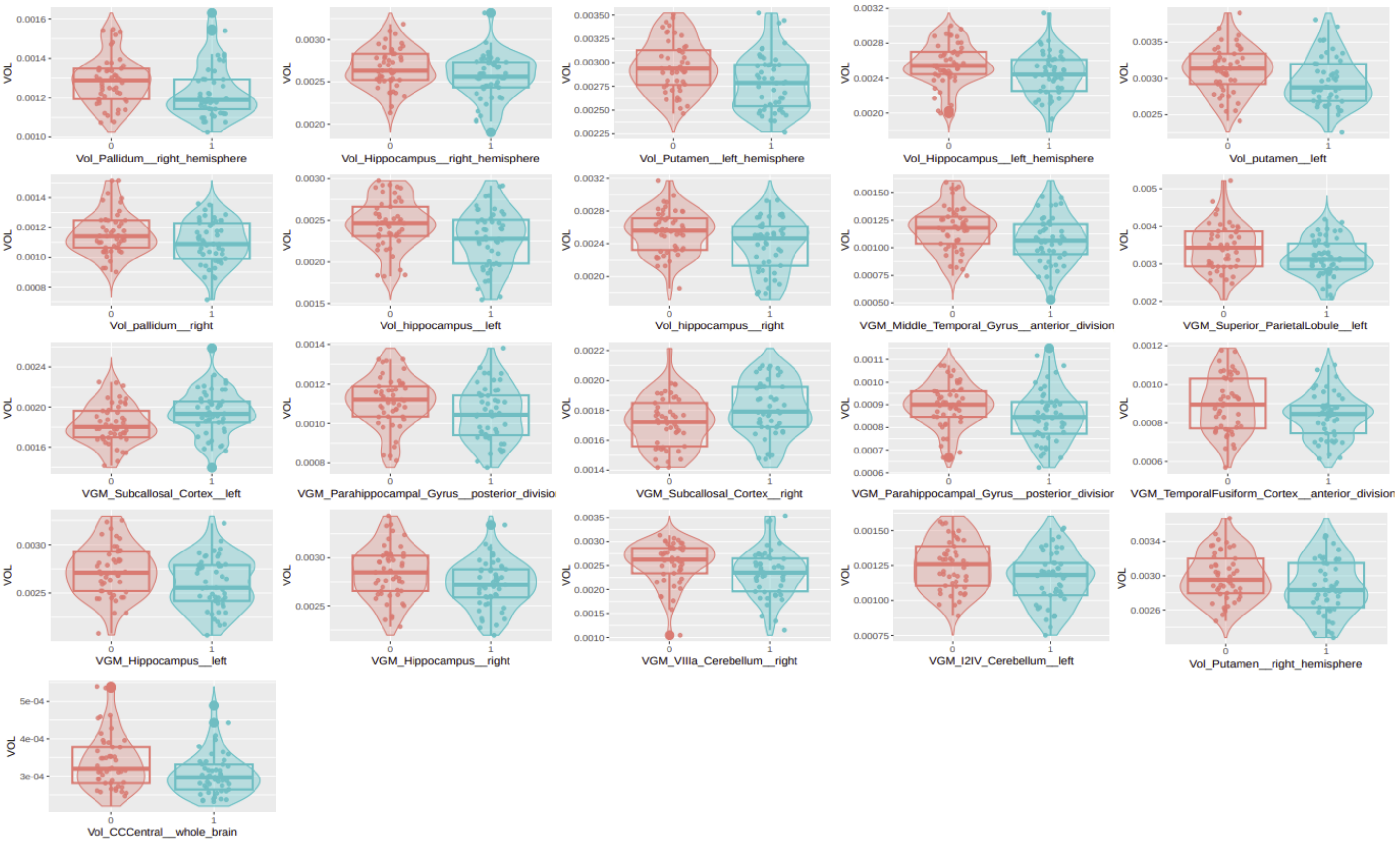


**Figure S7. Boxplots of cortical and subcortical volume differences between ET patients and low and high risk healthy controls**

Boxplots of cortical and subcortical volume differences between (1:1) matched ET patients and healthy controls (HC) (N:49) reveal regions normally associated with ET pathophysiology. These differences are more prominent in the ET vs low risk (LR) PRS group while fewer differences were found in the ET vs HR PRS group. (A) Depicts differences between the high risk group and patients. (B) Depicts differences between the low risk group and patients.
